# Gambling disorder symptom severity and mental health: an item-response-theory analysis of DSM-5 criteria for gambling disorder

**DOI:** 10.64898/2026.08.12.26360267

**Authors:** Franziska Vogl, Hans-Georg Wolff, Sven Buth, Jan Peters

**Affiliations:** Department of Psychology, Social Psychology, University of Cologne, Cologne, Germany; Department of Psychology, Biological Psychology, University of Cologne, Cologne, Germany; Department of Psychology, Organizational and Economic Psychology, University of Cologne, Cologne, Germany; Institute for Interdisciplinary Addiction and Drug Research (ISD), Hamburg, Germany

## Abstract

The present study examined the relationship between the DSM-5 diagnostic criteria for gambling disorder (GD) and gambling severity via an item response theory (IRT) analysis in two large German population survey data sets (Buth et al. (2022, 2024)). IRT-based person fit analyses may reveal atypical response patterns (e.g. endorsing criteria linked to higher levels of disorder severity, but not criteria linked to lower levels). We examined the link of such atypical response patterns and mental health as measured by the MHI-5, employing a 2-parameter-logistic (2PL) IRT model and a linear mixed model with random intercepts. Results largely replicated previously reported item severity rankings across both samples: GD criteria such as loss chasing and a preoccupation with gambling were generally linked to lower severity levels, whereas criteria such as withdrawal symptoms or job/family problems where generally linked to higher severity levels. Modelling revealed a reduced assessment sensitivity in lower gambling severity ranges. Furthermore, person fit analyses suggest that atypical symptom patterns may be linked to poorer mental health (MHI-5). Implications for the interpretability of total scores of endorsed criteria and the validity of diagnostic practices determining eligibility for treatment and financial compensation are discussed.

## Introduction

In recent years, studies have reported an increasing prevalence of gambling behaviour among adults and adolescents. In a review and meta-analysis of studies published between 2010 and 2024, Tran et al. (2024) estimated a global rate of gambling participation of 46.2 % among adults and 17.9 % among minors. This notion is corroborated by a representative German population survey by Buth et al. (2026) which recorded a gambling participation prevalence rate among adults of 36.4 % in 2025. Furthermore, growing gambling revenues have fuelled the rapid expansion of online gambling reaching increasingly young audiences (Montiel et al., 2021).

Excessive gambling can lead to negative consequences for individuals, those around them, and society at large. This includes not only detrimental effects on mental and physical health, but also financial hardship, unemployment, and the deterioration of personal relationships (Wardle et al., 2024). In severe cases, gambling has been linked to domestic violence, criminal behaviour and suicidality (Chui et al., 2018; Wardle et al., 2024). Furthermore rising criminality rates, personal indebtedness and treatment demands pose a substantial financial burden to society (National Research Council, 1999).

Negative consequences of gambling initially manifest at subclinical levels (problem gambling), and increase with increasing gambling disorder (GD) symptom severity (Ford & Håkansson, 2020). At the pathological level, GD is diagnosed if more than three out of a total of nine DSM-5 diagnostic criteria are endorsed, with four or five, six or seven and eight or nine endorsed criteria indicating a mild, moderate and severe disorder, respectively (see Table 1 for an overview of the GD DSM-5 criteria).

**Table 1.** DSM-5 diagnostic criteria for GD.

| No.<sup>1</sup> | Criterion | Description<sup>2</sup> | Endorsed<sup>3</sup> |  |
| --- | --- | --- | --- | --- |
|  |  |  | 2021 | 2023 |
| 1 | Preoccupation | Often preoccupied with gambling, e.g. persistent thoughts about gambling, planning next gambling experience, thinking of ways to earn money for gambling | 0.60 | 0.50 |
| 2 | Tolerance | Need to gamble with an increased amount of money to sustain the appeal or level of excitement | 0.42 | 0.34 |
| 3 | Cessation attempts<sup>4</sup> | Inability to decrease or quit gambling behaviour | 0.44 | 0.41 |
| 4 | Withdrawal | Withdrawal symptoms upon decreasing or quitting gambling, e.g. feeling agitated, restless, irritable | 0.21 | 0.17 |
| 5 | Escape | Gambling to escape personal problems or attenuate unpleasant emotions, e.g. fear or distress | 0.41 | 0.36 |
| 6 | Chasing losses<sup>5</sup> | Engaging in or increasing gambling behaviour to compensate for a monetary loss that has been caused by gambling | 0.53 | 0.47 |
| 7 | Concealment | Lying to others to conceal the extent of gambling behaviour | 0.33 | 0.26 |
| 8 | Jeopardizing | Jeopardizing relationships or missing out on events and opportunities in personal or work-related domains due to gambling | 0.22 | 0.17 |
| 9 | Bail-Out | Relying on others to cover debts or resolve financial problems caused by gambling | 0.18 | 0.15 |

*Note.* <sup>1</sup> The numbers of the diagnostic criteria match the numbers of items discussed in the analyses
<sup>2</sup> The descriptions are based on the diagnostic criteria for GD of the diagnostic and statistical manual of mental disorders (DSM-5; American Psychiatric Association, 2013) and reflect the questions of the screening employed by Buth et al. (2022; 2024)
<sup>3</sup> Proportion of participants who endorse the criterion in the 2021 and 2023 sample
<sup>4</sup> Referred to as “cessation” for short
<sup>5</sup> Referred to as “chasing” for short

In Germany, the GD diagnosis makes individuals eligible for psychotherapy financed by public health insurance. To ensure that gambling severity is accurately determined and help is afforded to those in need, DSM-5 GD criteria must function as a reliable and valid instrument. Two properties that are critical for this purpose are the spread of diagnostic criteria across the full range of gambling severity and the ability of each criterion to reliably discriminate between individuals above and below a given severity level. Criteria that cluster at a narrow severity range leave the instrument insensitive to GD outside that range, risking misclassifications that have direct consequences for treatment access. Criteria with poor discrimination add noise to the severity estimate and undermine diagnostic accuracy.

Item response theory (IRT) models directly assess these properties at the level of individual criteria (Reeve & Fayers, 2005). Within two-parameter-logistic (2PL) IRT models, the relationship between diagnostic criteria and the latent trait is described by item severity and item discrimination parameters (DeMars, 2010). The item severity parameter indicates the item’s location on the severity axis of the latent trait and is therefore informative on the spread of criteria along the gambling severity continuum. The item discrimination parameter marks the discriminatory propensity of the criteria.

Previous research consistently indicates that DSM-4 and DSM-5 criteria for GD differ in their association with gambling severity and in their diagnostic precision. Specifically, IRT analyses by Chui et al. (2018), Sleczka et al. (2015) and Strong & Kahler (2007) demonstrated that criteria vary in both their location along the severity continuum and their discrimination. While the three studies broadly agree in assigning similar sets of criteria to lower, middle and higher severity ranges, they are not fully consistent. The criteria tolerance, cessation and withdrawal fall into different severity categories across studies suggesting that the ordering of DSM-4 and DSM-5 criteria along the severity continuum remains to be established.

Studies employing other methodologies converge on a similar picture. Toce Gerstein et al. (2003) ordered DSM-4 criteria along a severity continuum in a manner largely consistent with previous IRT-based orderings (Chui et al., 2018; Sleczka et al., 2015; Strong & Kahler, 2007), while Stinchfield et al. (2005) found that DSM-4 criteria differ in discriminatory power and that diagnostic classification accuracy can be improved by weighting criteria accordingly. Orford et al. (2010) identified at least two DSM-4 criteria as performing poorly and Temcheff et al. (2016) found that among DSM-5 criteria, preoccupation best differentiated social from problem gamblers. In a network analysis of DSM-5 criteria, Lucas et al. (2024) identified withdrawal as the most central criterion, followed by tolerance and chasing, further underscoring that criteria are not equivalent in their relationship to gambling severity.

Taken together, the evidence suggests that DSM-5 criteria likely differ in the two properties established as critical, namely their spread across the severity continuum and their ability to discriminate between individuals differing in severity. The present study therefore applies IRT to an independent sample of individuals engaging in gambling to replicate and clarify the severity and discrimination estimates reported in previous studies, and to examine whether the two psychometric properties established as critical above are met by the DSM-5 criteria.

Relatedly, misclassification of individuals can also result from a mismatch between individual symptom patterns and the severity ordering of DSM-5 criteria. As the number of endorsed GD criteria is employed as an index of gambling severity in clinical practice, atypical disorder manifestations, i.e. endorsing criteria typically associated with higher severity levels (high criterion severity in IRT model), while failing to endorse criteria associated with lower severity levels (low criterion severity), may be classified as lower in severity (based on total criteria score) than endorsed criteria would indicate. It is therefore important to identify atypical disorder manifestations and investigate how they are represented by the DSM-5 GD criteria.

Findings by Challet-Bouju et al. (2026) underscore the notion of atypical disorder manifestations in GD. They identified four distinct clusters of individuals engaging in gambling that were characterized by different trajectories of DSM-5 criteria endorsement over time across the full spectrum of gambling involvement. Notably, similar total numbers of endorsed criteria were associated with different symptom configurations and temporal dynamics, particularly at subclinical levels. These findings suggest that individuals may deviate from the average symptom endorsement pattern implied by DSM-5 GD criteria, and that the total number of endorsed criteria may obscure meaningful heterogeneity in disorder manifestation and gambling severity.

The person-fit (PF) lends itself to the investigation of atypical symptom endorsement patterns as it quantifies the congruence between the manifestation of a respondent’s observed responses and IRT model predictions about the response pattern given that individual’s estimated level of the latent trait (Magis et al., 2012). In the context of psychodiagnostics, a high PF represents congruence between the disorder structure outlined by diagnostic criteria and the observed individual disorder manifestation. Low PF scores can be indicative of atypical symptom patterns or noise induced by random responding, measurement errors or unusual response styles (Magis et al., 2012; Tendeiro et al., 2016).

Other works have highlighted the relevance of including measures of PF in psychometric assessments. Conjin et al. (2015) reported that person misfit was related to the severity of psychological distress on the Outcome Questionnaire-45 (OQ-45) (Lambert et al., 2004) and Conrad et al. (2010) applied IRT-based PF analysis to identify unexpected psychiatric response patterns in persons with suicidal ideation.

In terms of diagnostic accuracy, low PF scores present an issue whenever they reflect atypical symptom patterns. Then, individual manifestations of gambling might yield an identical total number of endorsed criteria, despite being associated with different levels of psychological distress raising concerns about the validity of the total number of criteria as an index for gambling severity. If atypical symptom patterns involving criteria associated with higher severity, are in fact associated with greater psychological distress than typical symptom patterns involving criteria associated with lower severity, this should manifest as a relationship between PF and mental health within groups of identical total criteria count. An analysis of low PF and the relationship between an individual’s mental health and total criteria score is therefore warranted.

The aim of the present study is twofold. First, we examine the severity and discrimination of DSM-5 GD criteria in two independent, large-scale samples (the 2021 and 2023 gambling surveys conducted in Germany (Buth et al., 2022, 2024)) and compare estimates across samples and against the benchmark provided by Sleczka et al. (2015). Although previous IRT analyses have established that criteria differ in severity and discrimination, inconsistencies in criterion ordering across studies remain unresolved. Second, we employ a PF analysis to examine individual variability in criterion endorsement patterns. Specifically, we investigate the prevalence of atypical disorder manifestations and whether low PF scores indicative of unexpected response patterns are associated with poorer mental health outcomes. The inclusion of a PF analysis extends the methodology of previous IRT analyses of DSM-5 GD criteria.

We examine three hypotheses. We expect the estimates of criterion severity and discrimination parameters to differ among criteria indicating an average criteria endorsement pattern and differential functioning along the gambling severity continuum (1). Although random response patterns, unusual response styles and cognitive processing biases cannot be ruled out, we consider a low PF score to mainly reflect participants who endorse criteria linked to higher gambling severity while failing to endorse criteria associated with lower severity. Consequently, we expect lower PF scores to be linked to lower mental health, if the total number of endorsed criteria is held constant (2). We expect results to replicate in the second sample (3).

## Methods

### Participants

The present study constitutes a re-analysis of data from two separate large-scale surveys of gambling behaviour in Germany conducted by the Institute for Interdisciplinary Addiction and Drug Research in Hamburg (Institut für Interdisziplinäre Sucht- und Drogenforschung, ISD Hamburg) and the University of Bremen in 2021 (Buth et al., 2022) and 2023 (Buth et al., 2024). Researchers collected data on gambling behaviour and mental health among individuals aged 16 to 70 in the German population following a mixed-mode design, that combines telephone interviews and online questionnaires administered by the market research institute INFO. Data collection occurred from the 03.08.2021 to the 16.10.2021 and from the 01.08.2023 to the 16.10.2023 for the 2021 and 2023 surveys, respectively. In 2021, INFO commissioned Dynata and GapFish to recruit participants from online forums to increase the number of participants displaying more severe gambling behaviour. Buth et al. (2022) did not report these participants in the representative survey, but we included them in the present analysis.

Researchers informed participants that the survey contributed to a study about leisure-time behaviour and participants gave their informed consent (Meyer et al., 2024). Local legislation and institutional requirements required no ethical approval (Meyer et al., 2024). Meyer et al. (2024) described the exact data collection procedure.

In the present analyses, we included participants, who reported gambling at least once within the past 12 months (12-month-prevalence criterion) and who were above the age of eighteen. We excluded respondents fulfilling no diagnostic criteria for GD, as they do not inform the employed item-response theory analyses and could potentially cause an overestimation of the unidimensionality of the data (Sleczka et al., 2015). The total samples Buth et al. (2022, 2024) collected for the surveys in 2021 and 2023 consisted of N_1_ = 5318 and N_2_ = 4560 participants, respectively. In the first sample, n_1_ = 1917 participants fulfilled the inclusion criteria. After excluding one participant with missing data on the mental health variable, n_2_ = 979 participants of the second sample fulfilled the inclusion criteria. We performed all analyses using the 2021 sample (Buth et al., 2022) and examined whether the findings generalized to the 2023 sample (Buth et al., 2024). The 2021 sample comprises participants from the representatives survey (n_1a_ = 1070; Buth et al., 2022) and participants recruited from online forums (n_1b_ = 847).

### Preregistration

We preregistered the study on the Open Science Framework (https://osf.io/85zxc) prior to data analysis. For the assumption check we preregistered a scree plot and eigenvalue inspection for unidimensionality, Yen’s Q3 for local independence and plotting the ratio of people endorsing each criterion over the total sum of endorsed criteria for latent monotonicity. We preregistered the 2PL-IRT model for each of the two samples, the comparison of severity and discrimination parameter estimates between samples and to results by Sleczka et al. (2015), as well as the computation of the *lz\** index based on the two models. To assess whether the 2021 sample’s two recruitment strategies affected results, we compared 2PL-IRT models fitted separately for participants recruited through the representative survey by Buth et al. (2022) and participants recruited from online forums. This comparison was not preregistered. As an exploratory analysis not specified in the preregistration, we employed a linear mixed model to examine the relationship between the lz* index and the MHI-5 score.

### Measures

The present study focused on diagnostic criteria for GD and an estimate of mental health. Researchers assessed DSM-5 criteria for GD using a 17-item instrument derived from Stinchfield’s (2002) operationalization of DSM-4 criteria for pathological gambling, which Sleczka et al. (2015) adapted to DSM-5 following the removal of the illegal acts criterion. The instrument assesses each of the nine DSM-5 criteria with two items, except for one criterion, which is assessed by a single item. We considered a criterion as fulfilled if participants endorsed at least one of its pertaining items (Buth et al., 2022, 2024).

We employed the five-item Mental Health Inventory (MHI-5) as a rough screening index of mental health (Berwick et al., 1991; Buth et al., 2022, 2024). It reliably flags mood and anxiety disorders (Ten Have et al., 2024), but shows low sensitivity and specificity for somatoform and substance use disorders (Rumpf et al., 2001). We standardized the sum score to a range from 0-100 with higher values indicating better mental health.

### Model-agnostic analyses

To test for associations between MHI-5 scores and problem gambling symptom severity (no. of endorsed criteria), we examined scatter plots and tested for a negative association between both variables as reported previously (Browne et al., 2016; Moreira et al., 2023; Tran et al., 2024; Wardle et al., 2024). We calculated Spearman correlations, as the data did not fulfil the normality assumption for the variable of total endorsed criteria.

### Model-based analyses

#### Item Response Theory (IRT) Analysis

We based all analyses on item response theory (IRT) models fitted to the GD diagnostic criteria data, separately for each sample. In a preregistered analysis, we tested assumptions of unidimensionality and local independence separately for each sample using exploratory factor analysis (scree plot and eigenvalues) and Yen’s Q3, respectively. We examined latent monotonicity using the Mokken test, with monotonicity plots depicting endorsement proportions conditional on the total rest score inspected as preregistered diagnostic support.

As preregistered, we modelled the relationship between problem gambling severity and endorsement of the DSM-5 gambling criteria via a two-parameter-logistic (2PL) IRT model. Consistent with prior research (Chui et al., 2018; Sleczka et al., 2015; Strong & Kahler, 2007), we expected item discriminations to differ and therefore, freely estimated the discrimination parameter. Model implementation used hierarchical Bayesian parameter estimation following the documentation of IRT models in the Stan User’s Guide (Stan Development Team, n.d.). The probability *y* of participant *i* endorsing item *k* was modelled as

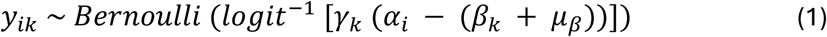

We modelled participants’ responses to items (*y*_ik_) as Bernoulli-distributed random variables, where the endorsement probability was given by the inverse logistic function of the difference between participant *i*’s latent trait α_i_, and item *k*’s criterion severity (*β*_k_ + *μ*_β_), scaled by the item discrimination parameter γ_k_.

Prior to implementation of the model in Stan, we fitted a 2PL-IRT model via *mirt* in R to assess model adequacy, inspecting RMSEA, TLI, CFA, M2, S-*χ*2 and infit/outfit statistics. Then, we estimated the final model (logit link) in Stan via *rstan* (Stan Development Team, 2025) using four chains (4,000 iterations; 2,000 warmup), yielding 8,000 post-warmup draws (see supplementary Figure 6; preregistered: https://osf.io/85zxc). We evaluated convergence by ensuring *R^* < 1.05 (Luo & Jiao, 2018), and extracted posterior medians and credible intervals. We inspected item characteristic curves (ICCs) to examine the probability of endorsing each item across levels of gambling severity. To verify the plausibility of participant-level parameter estimates we drew scatter plots and calculated Spearman correlations between posterior medians of individual participant gambling severity estimates α_j_ and the number of endorsed criteria.

To rule out potential differences between participants of the 2021 sample that we included within the representative survey by Buth et al. (2022) and those that were recruited via online forums, we fitted and compared separate IRT models for each subgroup of the 2021 sample in an exploratory analysis.

#### Person-Fit Analysis

In a preregistered PF analysis, we examined the association of participants’ deviation in response patterns from model predictions and mental health (MHI-5) employing two separate PF indices, the *lz\** PF index (Snijders, 2001) and the residual-based PF index. Here we focus on the *lz\** PF index, as it bears several advantages over the residual-based PF index. First, it accounts for participants’ complete response pattern through the joint likelihood of individual responses, in contrast to the residual-based PF index, which is based on summed item-level response probabilities (Tendeiro et al., 2016). Second, the performance of the *lz\** PF index and its predecessor *lz* (Drasgow et al., 1985) is empirically supported by simulation studies (Drasgow et al., 1987; Mousavi et al., 2016) and it is sensitive to spuriously low response vectors (Li & Olejnik, 1997; Nering & Meijer, 1998), which are relevant to the objective of this PF analysis.

Snijders (2001) first introduced the *lz\** PF index, which can be estimated via the *PerFit* R package by Tendeiro et al. (2016). A lower *lz\** value is associated with an increased deviation of the response patterns from model predictions. Following established practice, we classified scores below the cutoff of −1.645 as misfit (corresponding to an alpha level of α ∼ 0.05; Magis et al., 2012). We assessed mental health using the five-item MHI-5 (Berwick et al., 1991) which we standardized to a 0-100 scale.

To test whether the deviation of participants’ response patterns from model predictions (*lz\**) is associated with MHI-5 scores, we used two approaches. As variability of model-derived PF indices for participants fulfilling all nine diagnostic criteria is solely attributable to estimation noise, we excluded participants fulfilling all nine criteria from all PF analyses. First, in a preregistered analysis, we divided participants into *g* = 8 groups according to their total number of endorsed criteria (“total score groups”). Within these groups, we inspected scatter plots and calculated Spearman correlations between the standardized MHI-5 score and *lz\** PF index. Second, in a non-preregistered exploratory analysis, we assessed the effect of PF on mental health via a cross-classified linear mixed model with random intercepts for total score groups and sample. We performed this analysis on the combined dataset across both samples. Specifically, we modelled MHI-5 scores for participant *i* as

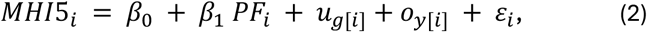

where the standardized mental health score for the *i*^th^ participant (*MHI*5_i_) is predicted by a fixed intercept (*β*_0_), a fixed slope (*β*_1_) multiplied by the PF of the *i*^th^ participant (*PF*_i_), a random intercept for the *i*^th^ participant in the *g*^th^ group (*u*_g[i]_) and in the *y*^th^ sample (*o*_y[i]_) as well as the error for the *i*^th^ participant (s_i_).

We calculated *lz\** PF scores using the *PerFit* R package (Tendeiro et al., 2016). The coefficient *β*_1_ represents the expected change in *MHI*5_i_ associated with a one-unit increase in *lz\**, holding total-score group and sample constant. A model including random slopes did not significantly improve model fit. Therefore, we retained no random slopes in the final model. We estimated the random intercept model by REML utilizing R packages *lme4* and *lmerTest*.

#### Software

We conducted all statistical analyses using the open-source software R version 4.5.2 (2025-10-31 ucrt) (R Core Team, 2025) and the Stan software for Bayesian data analysis v2.32.2 (Stan Development Team, 2023). We conducted the data preparation and PF analysis in RStudio working with posterior sample estimates of the 2PL-IRT models, which we estimated in Stan. The following R packages were used: bayesplot v. 1.13.0 (Gabry et al., 2019; Gabry & Mahr, 2025), car v. 3.1.3 (Fox & Weisberg, 2019), e 1071 v. 1.7.16 (Meyer et al., 2025), emmeans v. 1.11.2 (Lenth & Piaskowski, 2025), flextable v. 0.9.9 (Gohel & Skintzos, 2025), ggmirt v. 0.1.0 (Masur, 2025), glue v. 1.8.0 (Hester & Bryan, 2024), gt v. 1.0.0 (Iannone et al., 2025), gtsummary v. 2.3.0 (Sjoberg et al., 2021), knitr v. 1.50 (Xie, 2014, 2015, 2025), lme4 v. 1.1.37 (Bates et al., 2015), lmerTest v. 3.1.3 (Kuznetsova et al., 2017), lmtest v. 0.9.40 (Zeileis & Hothorn, 2002), mirt v. 1.44.0 (Chalmers, 2012a), mokken v. 3.1.2 (Ark, 2007, 2012), naniar v. 1.1.0 (Tierney & Cook, 2023), patchwork v. 1.3.2 (Pedersen, 2025), PerFit v. 1.4.7 (Tendeiro et al., 2016), psych v. 2.5.6 (Revelle, 2025), RColorBrewer v. 1.1.3 (Neuwirth, 2022), REdaS v. 0.9.4 (Maier, 2022), reshape2 v. 1.4.4 (Wickham, 2007), rmarkdown v. 2.29 (Allaire et al., 2024; Xie et al., 2018, 2020), rstan v. 2.32.7 (Stan Development Team, 2025), svglite v. 2.2.1 (Wickham et al., 2025), tidyverse v. 2.0.0 (Wickham et al., 2019).

## Results

We conducted all analyses in the 2021 sample and attempted to replicate the results in the 2023 sample. Henceforth, we will refer to the samples as the 2021 sample and the 2023 sample.

### Measurements

According to the DSM-5, patients get a GD diagnosis if they endorse four or more diagnostic criteria, with a mild, moderate and severe GD being indicated by the endorsement of four to five, six to seven and eight to nine criteria, respectively (American Psychiatric Association, 2013). Table 1 provides an overview of the nine diagnostic criteria (i.e., the nine items analysed below) and their percentage of endorsement in both samples. In total, n_1,GD_ = 735 (∼ 38.3 %) and n_2,GD_ = 276 (∼ 28.2 %) participants exceed the diagnostic threshold for GD in the 2021 and 2023 sample, respectively (see Table 4 in supplementary materials for a summary of GD prevalence).

### Model-agnostic analyses

To examine whether the MHI-5 score negatively correlates with the total number of endorsed criteria, as suggested by the relationship between gambling severity and a deterioration of mental health (Mide et al., 2023), we drafted scatter plots between variables (Figure 1). As the data did not meet normality and homoscedasticity assumptions, we employed a non-parametric correlation. The Spearman correlation between total endorsed criteria and MHI-5 score yielded a correlation coefficient of ρ ∼ −0.28 (p < 0.001) in both samples, indicating a medium effect size (Cohen, 1988).

**Figure 1.**
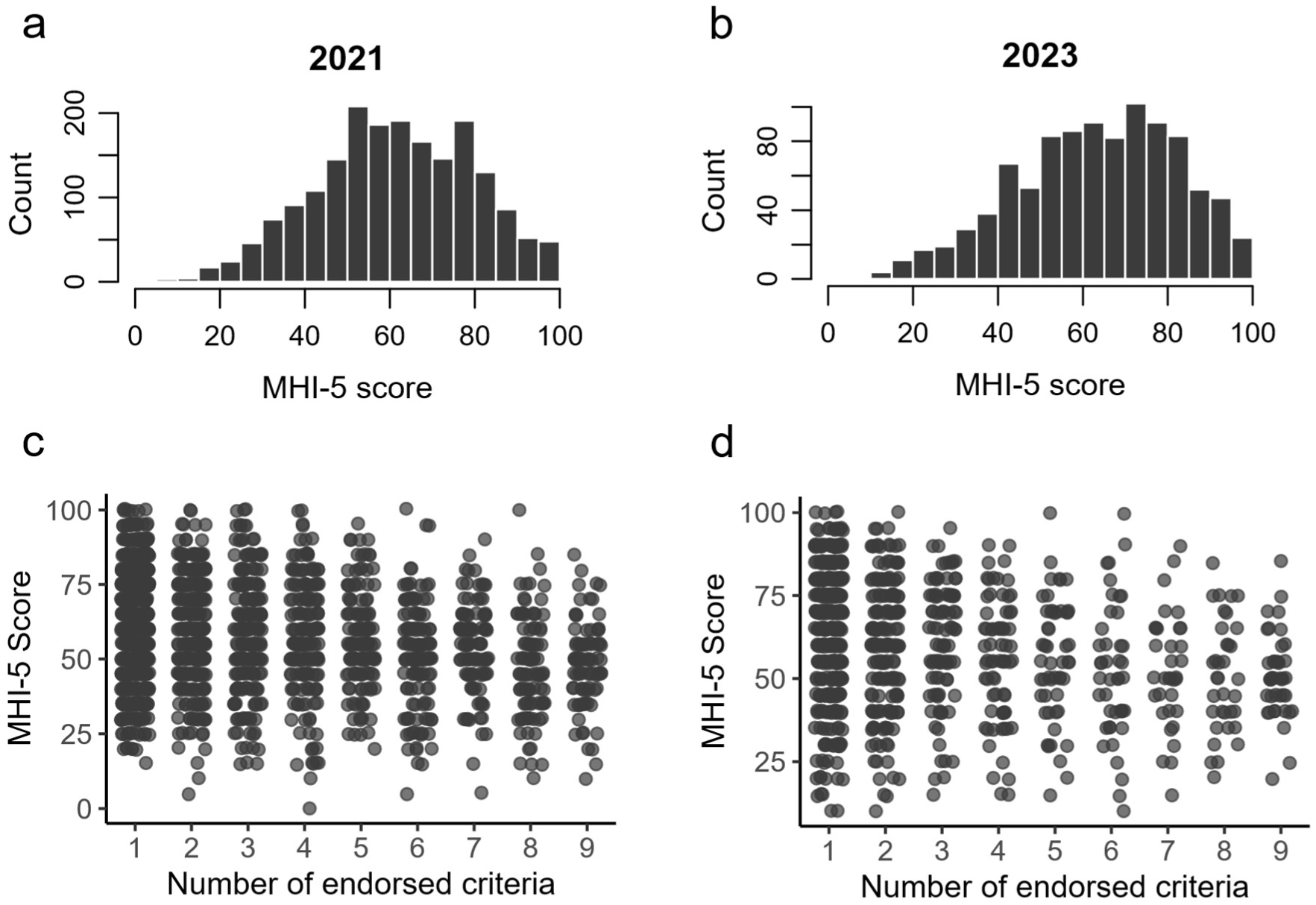
GD criteria and MHI-5 **a** and **b**, histograms of the standardized MHI-5 score for the 2021 and 2023 sample. **c** and **d**, scatter plots between the standardized MHI-5 score and the total number of endorsed criteria for the 2021 and 2023 sample. We jittered data points by 0.25 units horizontally and 0.40 units vertically.

### Model-based analyses

Exploratory factor analyses, Yen’s Q3 (see supplementary Table 5) and the Mokken test (no Crit values > 0; Ark, 2007; Mokken, 1971) confirmed assumptions of unidimensionality, local independence and latent monotonicity. Scree plots and factor loadings further supported a one-factor structure (see supplementary Figure 7 and Table 6).

### Item Response Theory (IRT)

To assess model adequacy, we fitted 2PL-IRT models to the 2021 and 2023 sample using the *mirt* package in R (Chalmers, 2012b). Several model fit indices suggest a good fit of the 2PL-IRT model for the 2021 sample (RMSEA ∼ 0.04, TLI ∼ 0.98 and CFI ∼ 0.99) and the 2023 sample (RMSEA ∼ 0.03, TLI ∼ 0.99 and CFI ∼ 0.99; Cai et al., 2023; Embretson & Reise, 2025). By contrast, the M2 statistic is significant in both samples (M2_21_ ∼ 107.99; df_21_ = 27; p_21_ < 0.001; M2_23_ ∼ 53.2; df_23_ = 27; p_23_ < 0.05) indicating a significant difference between the model and the data (Embretson & Reise, 2025).

We assessed item fit consulting infit, outfit and the S-*χ*^2^ statistic (Orlando & Thissen, 2000). All items showed non-standardized infit and outfit values within the conventional 0.5 – 1.5 range (see supplementary Table 7). By contrast, S-*χ*^2^ indicated acceptable fit only for items 2, 5 and 6 in the 2021 sample and items 2, 3, 4 and 6 in the 2023 sample.

Upon fitting 2PL-IRT models in Stan, we checked convergence by inspecting *R^* (Gelman et al., 2013). None of the estimates exceeded the critical value of 1.05 (Luo & Jiao, 2018). We extracted the medians and 95% credible intervals of the estimated item parameter distributions (see supplementary materials Table 8). To evaluate the plausibility of participants’ estimated severity (alpha parameter), we visualized and examined median severity estimates in relation to the observed number of endorsed criteria (Figure 2). We expected plausible estimates to reflect the right skewed distribution of participants’ total counts of endorsed criteria (supplementary Figure 8) and increase with endorsement counts. Spearman correlations between endorsed criteria and severity estimates were high (ρ ∼ 0.98, p < 0.001) in both the 2021 and 2023 samples.

**Figure 2.**
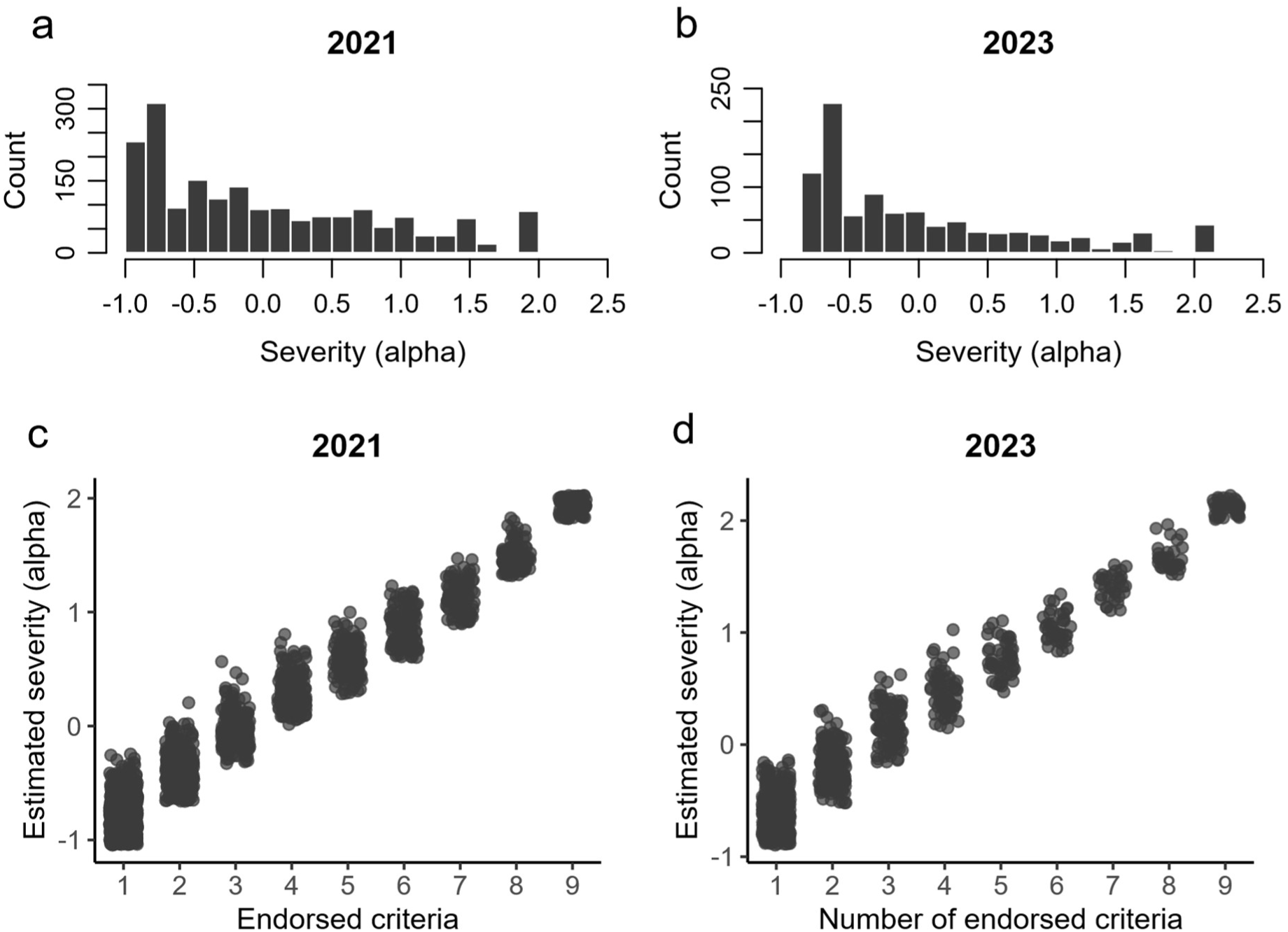
Alpha medians and endorsed criteria, **a** and **b** histograms of participants’ median severity estimates (alpha) from the 2PL-IRT models of the 2021 (**a**) and 2023 (**b**) samples. **c** and **d**, scatter plots between total counts of endorsed criteria and median severity estimates (alpha) for the 2021 (**a**) and 2023 (**b**) samples. We jittered data points by 0.25 units horizontally and 0.1 units vertically

Figure 3 displays the ICCs of the diagnostic criteria (see supplementary Figure 9 for total information functions (TIF)). As portrayed by the Kernel density plots, the overlap between the severity ranges covered by diagnostic criteria and respondents’ estimated gambling severity is moderate (Figure 3). This pattern is largely attributable to the comparatively high proportion of respondents with low levels of gambling severity.

**Figure 3.**
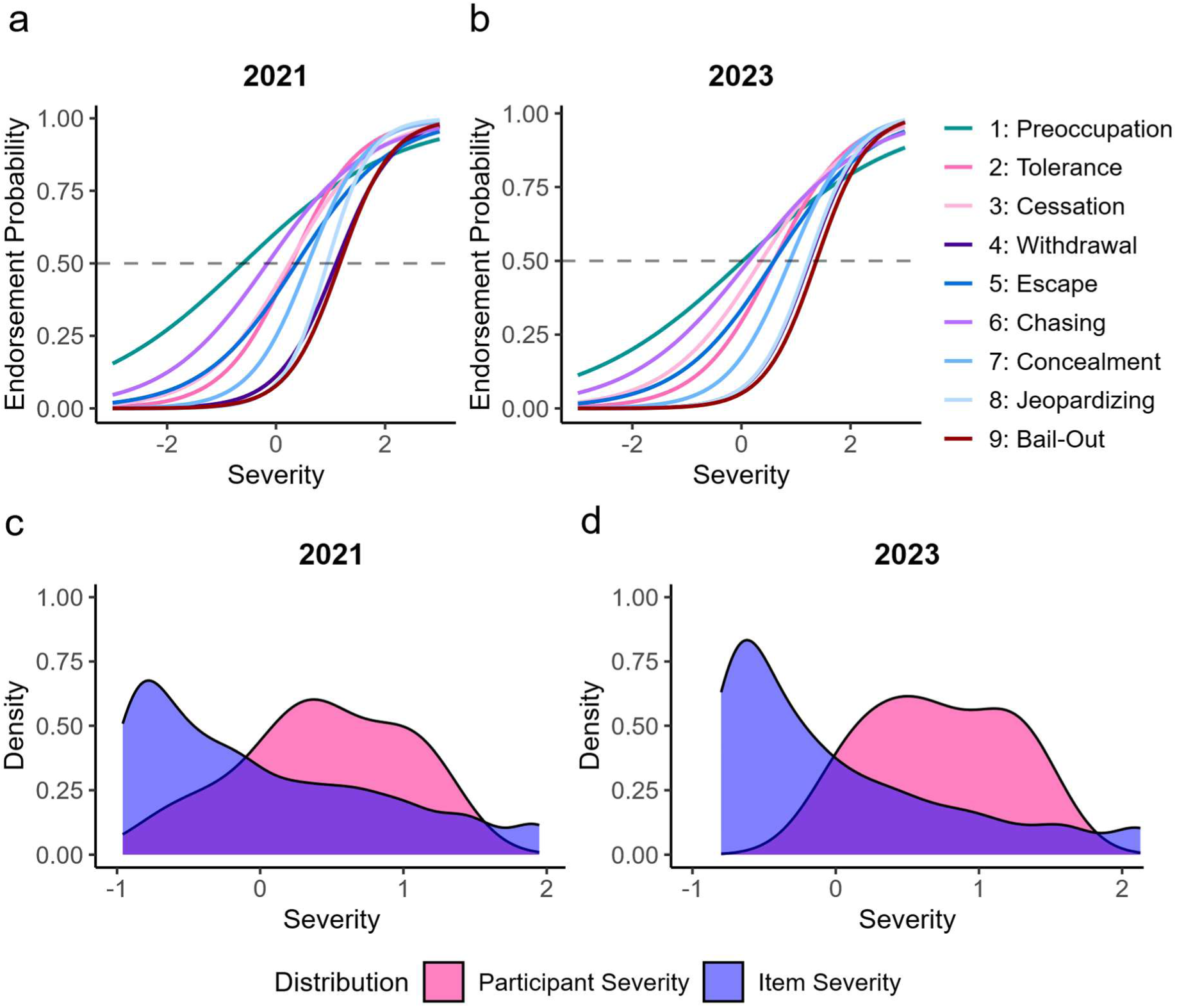
Item characteristic curves for the 2PL-IRT model of the 2021 sample (**a**) and the 2023 sample (**b**), Kernel density plots for item severity and participant severity estimates of the 2021 (**c**) and 2023 (**d**) sample

Item severity estimates indicate a criteria endorsement pattern by ranking them from lowest to highest. The item severity ranking in the 2023 sample fully replicated that of the 2021 sample, whereas the ranking based on the results by Sleczka et al. (2015) reversed the order of the escape of concealment criteria, constituting a partial replication (Table 2). The ranking of the item discrimination estimates of the 2023 sample fully replicated the ranking of the 2021 sample, as well (Table 2). Results from the present analysis largely replicate the item discrimination rankings reported by Sleczka et al. (2015).

**Table 2.** Comparison of the item severity (a) and discrimination (b) ranking between the 2021 and 2023 samples and results by Sleczka et al. (2015)

| Item severity (beta) |  |  |  |  |  |  |
| --- | --- | --- | --- | --- | --- | --- |
| 2021 |  |  | 2023 |  | Slecza et al. (2015) |  |
| Rank<sup>1</sup> | Estimate | Criterion | Estimate | Criterion | Estimate | Criterion |
| 1 | -0.61 | Preoccupation | 0.03 | Preoccupation | -0.64 | Preoccupation |
| 2 | -0.16 | Chasing losses | 0.15 | Chasing losses | -0.28 | Chasing losses |
| 3 | 0.24 | Cessation | 0.37 | Cessation | 0.48 | Cessation |
| 4 | 0.28 | Tolerance | 0.60 | Tolerance | 0.51 | Tolerance |
| 5 | 0.38 | Escape | 0.60 | Escape | 0.80 | Concealment |
| 6 | 0.59 | Concealment | 0.89 | Concealment | 1.28 | Escape |
| 7 | 0.94 | Jeopardizing | 1.23 | Jeopardizing | 1.32 | Jeopardizing |
| 8 | 1.10 | Withdrawal | 1.25 | Withdrawal | 1.35 | Withdrawal |
| 9 | 1.17 | Bail-Out | 1.37 | Bail-Out | 1.62 | Bail-Out |

*Note.* <sup>1</sup> Criteria are ordered by their estimated item severity from lowest to highest

**b**
| Item discrimination (gamma) |  |  |  |  |  |  |
| --- | --- | --- | --- | --- | --- | --- |
| 2021 |  |  | 2023 |  | Slecza et al. (2015) |  |
| Rank<sup>1</sup> | Estimate | Criterion | Estimate | Criterion | Estimate | Criterion |
| 1 | 0.71 | Preoccupation | 0.68 | Preoccupation | 0.51 | Chasing losses |
| 2 | 1.06 | Chasing losses | 0.93 | Chasing losses | 0.53 | Preoccupation |
| 3 | 1.16 | Escape | 1.14 | Escape | 0.74 | Escape |
| 4 | 1.32 | Cessation | 1.16 | Cessation | 1.04 | Cessation |
| 5 | 1.61 | Tolerance | 1.46 | Tolerance | 1.05 | Concealment |
| 6 | 1.89 | Concealment | 1.76 | Concealment | 1.08 | Tolerance |
| 7 | 1.89 | Withdrawal | 2.11 | Withdrawal | 1.62 | Jeopardizing |
| 8 | 2.11 | Bail-Out | 2.13 | Bail-Out | 2.01 | Withdrawal |
| 9 | 2.52 | Jeopardizing | 2.13 | Jeopardizing | 2.56 | Bail-Out |

*Note.* <sup>1</sup> Criteria are ordered by their estimated item discrimination from lowest to highest

To investigate possible differences in criteria function between the subsamples of the 2021 sample, i.e. participants included in the representative survey by Buth et al. (2022) versus participants from online forums, we fitted the data of each subsample with a 2PL-IRT model. Figure 4 shows the ICCs and estimate rankings pertaining to each subsample. The item severity rankings differed in the ordering of two criterion pairs across subsamples, whereas the item discrimination rankings differed in the ordering of three criterion pairs. In general, criteria positions along the low-high continuum for criteria severity and discrimination estimates are comparable between subsamples (Figure 4).

**Figure 4.**
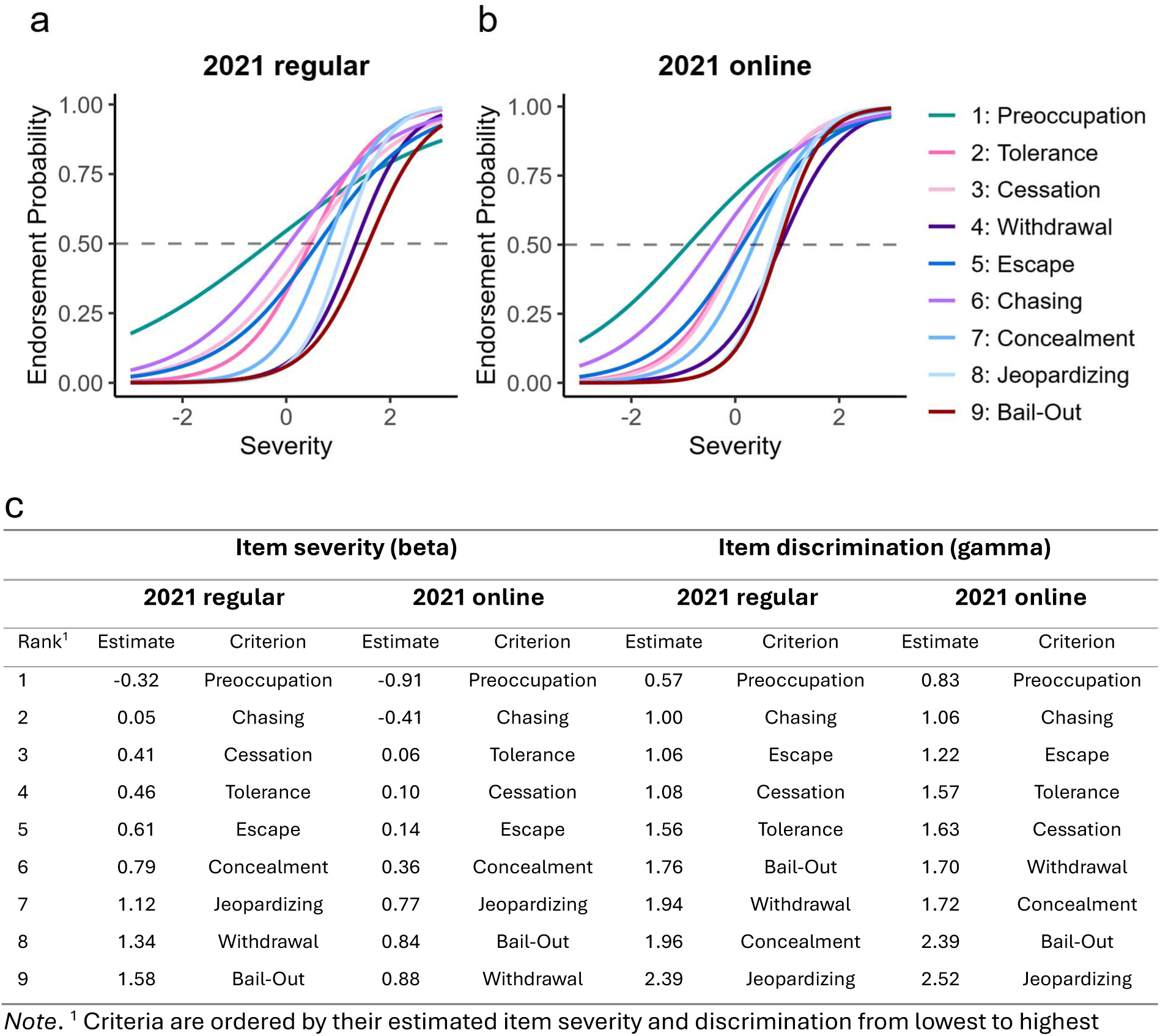
Comparison between the regular and online subset of the 2021 sample. ICCs of the regular (**a**) and the online subset (**b**), ranking of item severity and item discrimination estimates for each subset (**c**).

### Person-Fit Analysis

Comparing *lz\** scores to a predefined cut-off value of < −1.645, we flagged 109 respondents (∼ 5.7 %) and 44 respondents (∼ 4.5 %) as inconsistent with the IRT model of the 2021 and 2023 sample, respectively. The bar plots in Figure 5 display Spearman correlation estimates between *lz\** and MHI-5 scores within groups of total endorsed criteria counts. As smaller *lz\** scores correspond to deviating response patterns, we expected *lz\** scores to increase with increasing MHI-5 scores, i.e. positive correlation estimates. Notably, respondents of the 2021 sample endorsing two, six and eight criteria and respondents of the 2023 sample endorsing seven and eight criteria display correlations that counter theoretical predictions of directionality. In the 2021 sample, correlations reached statistical significance within both the group of respondents endorsing one criterion and the group endorsing eight criteria (ρ_lz,1_ ∼ 0.09, p_lz,1_ < 0.05; ρ_lz,8_ ∼ −0.24, p_lz,8_ < 0.05). In the 2023 sample, none of the correlations reached statistical significance. Correlations pertaining to the residual-based PF index are reported in the supplementary Figure 10.

**Figure 5.**
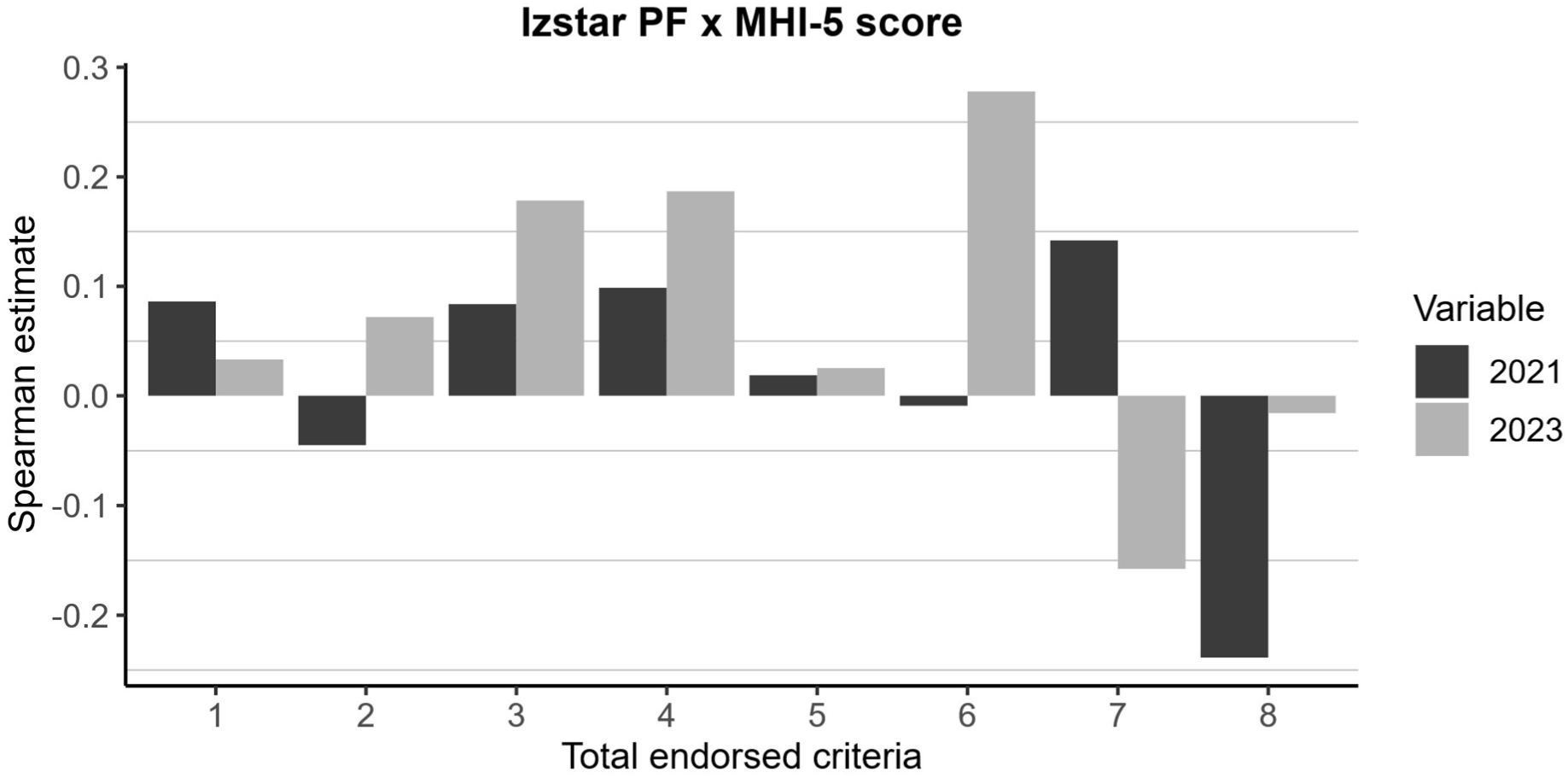
Correlations between *lz\** PF and MHI-5 scores for groups of total endorsed criteria of each sample

Researchers ran a random intercept model including the *lz\** PF index as predictor, the MHI-5 score as the outcome variable and random intercepts for the variables of groups based on total number of endorsed criteria and sample year (Table 3). We included no random slopes, as the *χ*^2^ test did not indicate a significantly better fit of the random slope model in comparison to random intercept model (*χ*^2^ ∼ 0.87; df = 4; p ∼ 0.93).

**Table 3.**
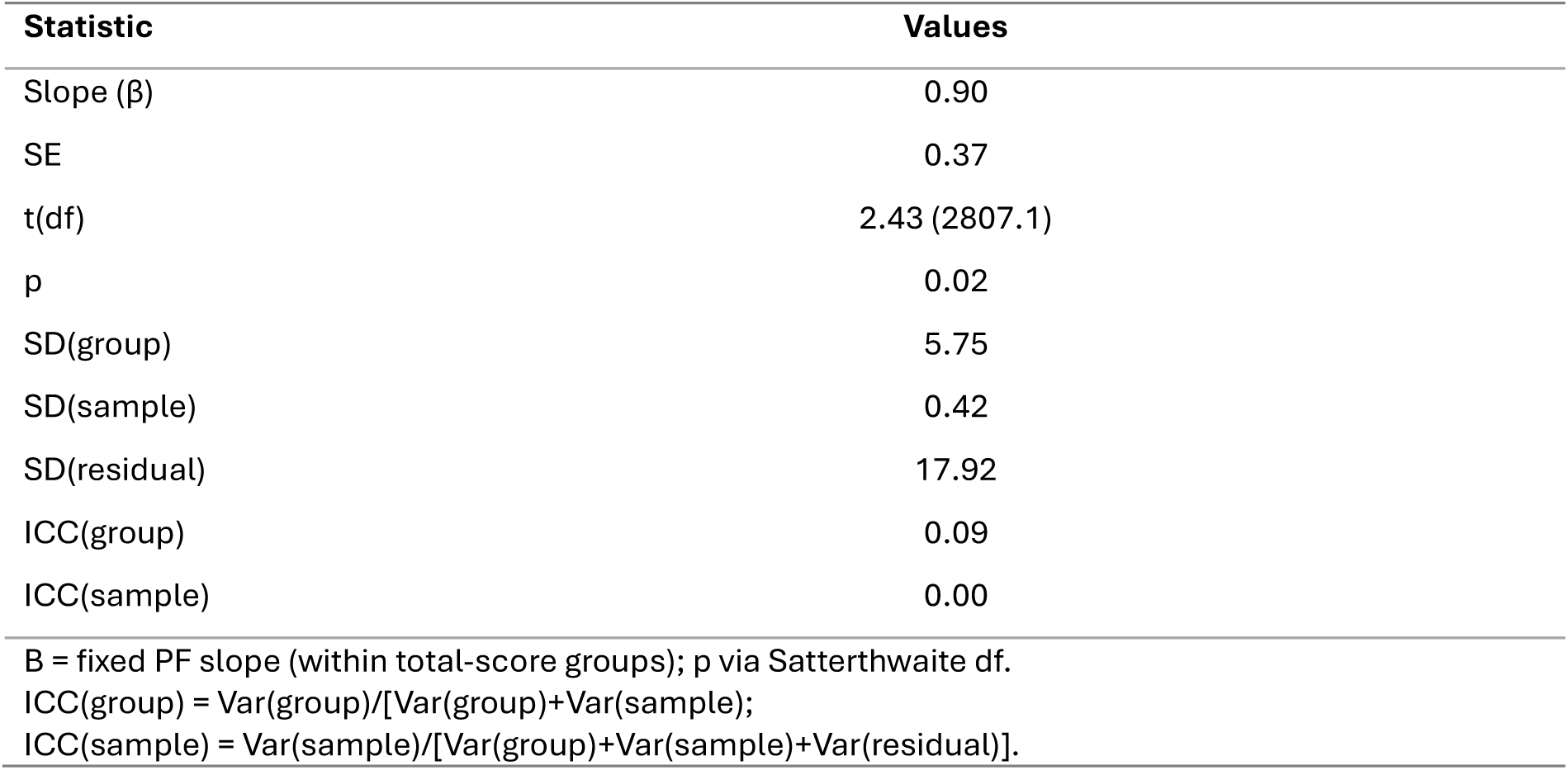
Random intercept model for lz* PF index.

| Statistic | Values |
| --- | --- |
| Slope ($\beta$) | 0.90 |
| SE | 0.37 |
| t(df) | 2.43 (2807.1) |
| p | 0.02 |
| SD(group) | 5.75 |
| SD(sample) | 0.42 |
| SD(residual) | 17.92 |
| ICC(group) | 0.09 |
| ICC(sample) | 0.00 |
| B = fixed PF slope (within total-score groups); p via Satterthwaite df.<br>ICC(group) = $\text{Var}(\text{group}) / [\text{Var}(\text{group}) + \text{Var}(\text{sample})]$;<br>ICC(sample) = $\text{Var}(\text{sample}) / [\text{Var}(\text{group}) + \text{Var}(\text{sample}) + \text{Var}(\text{residual})]$. | |

Holding the total number of endorsed criteria and sample year constant, the fixed effect of the *lz\** PF index on the mental health score was significant (β∼ 0.90, SE ∼ 0.37, p ∼ 0.02), which is in line with expectations. ICC values lie between within [0.00 – 0.09] indicating a low proportion of total variance explained by between-group and between-sample differences. Supplementary table 9 displays results of the random intercept model for the residual-based PF index.

## Discussion

Here we examined the structure of the DSM-5 criteria for GD in the data from two large general population surveys conducted by Buth et al. (2022, 2024) in 2021 and 2023 in Germany. We applied two parameter item response theory models (2PL-IRT) and hierarchical Bayesian parameter estimation.

IRT analyses revealed a specific criteria endorsement pattern and a systematic increase in discriminatory precision of diagnostic criteria associated with increasing levels of gambling severity. In other words, criteria signalling more severe problems were also the most informative ones. The results of the 2023 sample largely replicated findings in the 2021 sample, and both were comparable to results by Sleczka et al. (2015). We also found a negative relationship between the total number of endorsed criteria and mental health measured by the MHI-5 score (Berwick et al., 1991) in both samples. Together, these results confirm a characteristic overall criterion endorsement pattern linked to different symptoms. We next used person fit (PF) analyses to investigate atypical endorsement patterns (i.e. endorsing criteria linked to high severity, but failing to endorse criteria linked to low severity) and their effect on mental health outcomes (MHI-5). Correlational and exploratory PF analyses offered tentative support for a positive relationship between the respondents’ PF and mental health, when holding the total number of endorsed criteria constant.

### Model fit and plausibility

Given the positive relationship between medians of the estimated alpha parameter and the total score of endorsed criteria (Figure 2), we considered both 2PL-IRT models plausible. While most model fit statistics indicated excellent model fit, the M2 test suggested a discrepancy between model and observed data in both samples. As the test is known to be highly sensitive to even trivial deviations from model fit for large sample sizes, these results are not unexpected (Stone, 2021). Similarly, infit and outfit values were within the acceptable range (supplementary Table 7), whereas the S-*χ*2 test suggested poor fit for several items in both samples. Despite overall acceptable results, indications of possible misfit should be kept in mind when interpreting results.

### Model-agnostic analyses

Although no previous study has directly correlated the total number of endorsed DSM-5 criteria with MHI-5 scores, the present finding of a medium effect is broadly consistent with the established link between GD and poor mental health (Buth et al., 2017; Grant et al., 2017). Yet, it is smaller than the moderate-to-large effect sizes found between DSM-5 criterion counts and depression and anxiety scores measured with the PHQ and GAD-7 (Cabral et al., 2025). This may be attributable to limitations of the MHI-5, which is a coarse and brief self-report measure with only five items. Although it has been found to reliably flag psychological distress in relation to subclinical and clinical symptoms of anxiety and depression (Bültmann et al., 2006; Rivera-Riquelme et al., 2019; Strand et al., 2003; Ten Have et al., 2024), it may be a less robust measure in substance use disorders (Rumpf et al. 2001), and there is limited research validating the MHI-5 for behavioural addictions (independent of mood or anxiety disorders).

### Criteria endorsement pattern

Criterion severity and discrimination indices did differ across diagnostic criteria, revealing a stable endorsement pattern which we observed in both samples and which was largely consistent with the findings by Sleczka et al. (2015). Certain criteria, such as preoccupation with gambling and chasing losses, tended to be endorsed by individuals with comparatively mild gambling problems, whereas criteria such as jeopardizing relationships because of gambling or experiencing withdrawal symptoms when attempting to quit, were endorsed only among those with more severe gambling problems (Table 2). This ranking replicated exactly between samples. Compared to Sleczka et al. (2015), only the criteria escape and concealment were swapped, constituting a replication with minor deviations. These findings outline an approximate progression in how gambling disorder symptoms emerge as gambling severity increases.

In general, criteria’s assessment precision (discrimination) was higher for those criteria linked to higher gambling severity levels compared to those linked to lower levels. This order replicated exactly between samples (Table 2) and mirrors the trend reported by Sleczka et al. (2015). It is additionally reflected in higher test information for severity levels above the mean (supplementary Figure 9).

The results are consistent with Chui et al. (2018) and structural investigations of DSM-4 criteria for pathological gambling suggesting differential item functioning and an unequal diagnostic impact of diagnostic criteria (Orford et al., 2010; Stinchfield, 2002; Strong & Kahler, 2007; Toce-Gerstein et al., 2003). A recent network analysis of DSM-5 criteria for GD by Lucas et al. (2024) provides complementary evidence underscoring the centrality of specific diagnostic criteria, such as withdrawal, tolerance and chasing losses with an increased influence on GD severity and diagnostic weight. Taken together present IRT analyses reveal a reliable and generalizable structure of DSM-5 diagnostic criteria. In line with previous investigations, it supports the notion of unequal diagnostic impact of the criteria, demonstrating increased precision for higher levels of gambling behaviour.

### Person-fit analyses

PF and mental health were related within groups of endorsed criteria counts largely in the expected direction. Respondents with atypical response patterns tended to show poorer mental health than those with typical patterns with only a small number of groups deviating from this pattern. The pattern replicated descriptively across both samples. Results from the non-preregistered random intercept model further supported a negative effect of atypical disorder manifestations on respondents’ mental health when holding the total number of endorsed criteria constant. Most of the variability in mental health arose at the individual level rather than differing systematically between groups or samples. Yet, this effect remained small and provides only tentative support, suggesting that any underlying effect, if present, is likely small and difficult to detect with the present analytical approach. Ultimately, further investigations will be necessary to converge on an understanding of whether atypical disorder manifestations in GD present with more severe mental health problems compared to their typical counterparts. If future investigations supported this notion, this would raise concerns about the interpretability of the total count of endorsed criteria as an index of GD severity, given its role in the allocation of resources such as therapy and financial aid.

### Limitations

The 2021 sample was extended by the inclusion of individuals participating in gambling recruited via online forums. The IRT models based on participants included in the representative survey by Buth et al. (2022) versus participants from online forums were highly comparable in their ranking of criteria severity estimates and the systematic increase in discriminatory propensity with increasing criteria severity, suggesting a consistent structure of DSM-5 diagnostic criteria for GD among both 2021 subsamples. However, the 2021 and 2023 samples were not strictly comparable in sample size and composition, e.g. with respect to types of gambling.

The reduced power of correlational analyses caused by dividing respondents into groups based on the total number of endorsed criteria and the calculation of Spearman instead of Pearson correlation coefficients due to a violation of the normality assumption, is another limitation of the study. It may contribute to the lack of significant results of the preregistered correlational analyses (but not the exploratory linear mixed model). Furthermore, there are some indications for possible model misspecification and an item-person mismatch (Figure 9), which may introduce noise to PF estimates.

Concerns about the measures examined in the PF analysis also limit the precision and interpretability of results. While the MHI-5 may be limited in validity pertaining to psychological distress caused by gambling (Rumpf et al., 2001), the PF index *lz\** may be misaligned with the rationale of the present analysis. Specifically, IRT models estimate the probability of endorsing each criterion as a function of gambling severity, with ICCs providing a graphical representation of these endorsement probabilities. The *lz\** score quantifies the extent to which observed responses deviate from these probabilities. In the present analysis, deviation is defined relative to an average endorsement pattern reflected by the ranking of criterion severity estimates, such that a deviating response pattern (low PF) is characterised by endorsing criteria with higher severity estimates while failing to endorse criteria with lower severity. This definition holds as long as ICCs do not cross. When crossings occur, response patterns theoretically representing stronger deviations (average endorsement pattern) are mathematically captured as weaker deviations (endorsement probabilities), for respondents whose gambling severity falls above the crossing point.

This misalignment is inherent to model-based PF indices such as *lz*, *lz\** and *Zh* (Drasgow et al., 1987; Snijders, 2001), which derive expected response probabilities directly from the 2PL ICCs and therefore implicitly assume a person-location-dependent criterion ordering rather than a fixed one. Although nonparametric PF indices such as Guttman errors or U3 (Guttman, 1950; Van Der Flier, 1982) quantify PF relative to a fixed criteria ordering independent of model-based endorsement probabilities, they disregard variation in criterion discrimination. Given prior evidence that GD criteria differ in their discriminatory capacity (Chui et al., 2018; Sleczka et al., 2015), the present analysis used a 2PL-IRT model and the lz* index to retain information about both criterion severity and discrimination. The observed misalignment should therefore not be seen as an avoidable methodological error, but as a limitation arising from the tension between a simplified theoretical interpretation of criterion ordering and the full probabilistic structure of the 2PL-IRT model.

## Conclusion

To conclude, the findings indicate a stable relationship between the DSM-5 diagnostic criteria for GD and gambling as described by 2PL-IRT models with severity and discrimination parameters replicating between the 2021 and 2023 sample. Shortcomings of the DSM-5 criteria were identified pertaining to a reduced assessment sensitivity in lower gambling severity ranges. PF analyses suggested potential misclassifications of atypical symptom trajectories, that do not align with the average criteria endorsement pattern demonstrated by IRT analyses and show tentative association with decreased mental health outcomes. This raises concerns about the interpretability of total scores of endorsed criteria and the validity of diagnostic practices determining eligibility for treatment and financial compensation. Future studies might explore possible modifications of DSM-5 criteria for GD, e.g. the inclusion of criteria weights, to account for and accurately assess gambling severity in the case of atypical symptom trajectories.

## Data Availability

All data produced in the present study are available upon reasonable request to the authors.

## Author contributions

FV, SB and JP conceived the study. FV analyzed the data and wrote the paper. JP supervised the project. All authors provided revisions and theoretical input.

## Conflict of interest

FV, HGW, SV and JP have no conflict of interest to declare.

## Funding

This work was funded by the Biological Psychology Group of the University of Cologne.

## Notes

### Competing Interest Statement

The authors have declared no competing interest.

### Author Declarations

The Ethics Committee of the Hamburg Chamber of Physicians (Hamburger Aerztekammer) waived ethical approval for this work (reference number: 2023-300315-WF). This pertains to the survey data the present study uses for analysis.

## References

Allaire, J., Xie, Y., Dervieux, C., McPherson, J., Luraschi, J., Ushey, K., Atkins, A., Wickham, H., Cheng, J., Chang, W., & Iannone, R. (2024). rmarkdown: Dynamic documents for r. https://github.com/rstudio/rmarkdown

American Psychiatric Association. (2013). Diagnostic and Statistical Manual of Mental Disorders (Fifth Edition). American Psychiatric Association. 10.1176/appi.books.9780890425596

Ark, L. A. V. D. (2007). Mokken Scale Analysis in *R*. Journal of Statistical Software, 20(11). 10.18637/jss.v020.i11

Ark, L. A. V. D. (2012). New Developments in Mokken Scale Analysis in *R*. Journal of Statistical Software, 48(5). 10.18637/jss.v048.i05

Bates, D., Mächler, M., Bolker, B., & Walker, S. (2015). Fitting Linear Mixed-Effects Models Using **lme4**. Journal of Statistical Software, 67(1). 10.18637/jss.v067.i01

Berwick, D. M., Murphy, J. M., Goldman, P. A., Ware, J. E., Barsky, A. J., & Weinstein, M. C. (1991). Performance of a Five-Item Mental Health Screening Test: Medical Care, 29(2), 169–176. 10.1097/00005650-199102000-00008

Browne, M., Langham, E., Rawat, V., Greer, N., Li, E., Rose, J., Rockloff, M., Donaldson, P., Thorne, H., Goodwin, B., Bryden, G., & Best, T. (2016). Assessing gambling-related harm in Victoria: A public health perspective. Victorian Responsible Gambling Foundation.

Bültmann, U., Rugulies, R., Lund, T., Christensen, K. B., Labriola, M., & Burr, H. (2006). Depressive symptoms and the risk of long-term sickness absence: A prospective study among 4747 employees in Denmark. Social Psychiatry and Psychiatric Epidemiology, 41(11), 875–880. 10.1007/s00127-006-0110-y

Buth, S., Gerhard, M., Rosenkranz, M., & Kalke, J. (2024, March). Glücksspielteilnahme und glücksspielbezogene Probleme in der Bevölkerung—Ergebnisse des Glückspiel-Survey 2023. Institut für interdisziplinäre Sucht- und Drogenforschung (ISD).

Buth, S., Meyer, G., & Kalke, J. (2022, March). Glücksspielteilnahme und glücksspielbezogene Probleme in der Bevölkerung—Ergebnisse des Glückspiel-Survey 2021. Institut für interdisziplinäre Sucht- und Drogenforschung (ISD).

Buth, S., Meyer, G., Rosenkranz, M., & Kalke, J. (2026, March). Glücksspielteilnahme und glücksspielbezogene Probleme in der Bevölkerung—Ergebnisse des Glücksspiel-Survey 2025. Institut für interdisziplinäre Sucht- und Drogenforschung (ISD).

Buth, S., Wurst, F. M., Thon, N., Lahusen, H., & Kalke, J. (2017). Comparative Analysis of Potential Risk Factors for at-Risk Gambling, Problem Gambling and Gambling Disorder among Current Gamblers—Results of the Austrian Representative Survey 2015. Frontiers in Psychology, 8, 2188. 10.3389/fpsyg.2017.02188

Cabral, R. M., Cunha, C., Pires, P., Santos, I., Cunha, F., Oliveira, A., Gil, N., & Coias, F. (2025). The rise of online gambling addiction: A mental health challenge. European Psychiatry, 68(S1), S103–S104. 10.1192/j.eurpsy.2025.308

Cai, L., Chung, S. W., & Lee, T. (2023). Incremental Model Fit Assessment in the Case of Categorical Data: Tucker–Lewis Index for Item Response Theory Modeling. Prevention Science, 24(3), 455–466. 10.1007/s11121-021-01253-4

Challet-Bouju, G., Brault, V., Perrot, B., Jeu-Group, Desmée, S., & Grall-Bronnec, M. (2026). A clustering based on the dynamics of DSM-5 criteria for gambling disorder: A 5-year follow-up of gamblers with and without gambling disorder. Journal of Behavioral Addictions, 2006.2025.00099. 10.1556/2006.2025.00099

Chalmers, R. P. (2012a). **mirt**: A Multidimensional Item Response Theory Package for the *R* Environment. Journal of Statistical Software, 48(6). 10.18637/jss.v048.i06

Chalmers, R. P. (2012b). **mirt**: A Multidimensional Item Response Theory Package for the *R* Environment. Journal of Statistical Software, 48(6). 10.18637/jss.v048.i06

Chui, W.-Y., Lee, S.-K., Mok, Y.-L., & Tsang, C.-K. (2018). The Diagnostic Criteria of Gambling Disorder of DSM-5 in Chinese Culture: By Confirmatory Factor Analysis (CFA) and Item Response Theory (IRT). In M.-T. Leung & L.-M. Tan (Eds), Applied Psychology Readings (pp. 73–86). Springer Singapore. 10.1007/978-981-10-8034-0_5

Cohen, J. (1988). Statistical Power Analysis for the Behavioral Sciences (Second). Lawrence Erlbaum Associates.

Conijn, J. M., Emons, W. H. M., De Jong, K., & Sijtsma, K. (2015). Detecting and Explaining Aberrant Responding to the Outcome Questionnaire–45. Assessment, 22(4), 513–524. 10.1177/1073191114560882

Conrad, K. J., Bezruczko, N., Chan, Y.-F., Riley, B., Diamond, G., & Dennis, M. L. (2010). Screening for atypical suicide risk with person fit statistics among people presenting to alcohol and other drug treatment. Drug and Alcohol Dependence, 106(2–3), 92–100. 10.1016/j.drugalcdep.2009.07.023

DeMars, C. (2010). Item Response Theory. Oxford University Press. 10.1093/acprof:oso/9780195377033.001.0001

Drasgow, F., Levine, M. V., & McLaughlin, M. E. (1987). Detecting Inappropriate Test Scores with Optimal and Practical Appropriateness Indices. Applied Psychological Measurement, 11(1), 59–79. 10.1177/014662168701100105

Drasgow, F., Levine, M. V., & Williams, E. A. (1985). Appropriateness measurement with polychotomous item response models and standardized indices. British Journal of Mathematical and Statistical Psychology, 38(1), 67–86. 10.1111/j.2044-8317.1985.tb00817.x

Embretson, S. E., & Reise, S. P. (2025). Item Response Theory: Foundations for Psychologists and Social Scientists (2nd edn). Routledge. 10.4324/9781315726557

Ford, M., & Håkansson, A. (2020). Problem gambling, associations with comorbid health conditions, substance use, and behavioural addictions: Opportunities for pathways to treatment. PLOS ONE, 15(1), e0227644. 10.1371/journal.pone.0227644

Fox, J., & Weisberg, S. (2019). An R Companion to Applied Regression (Third). Sage. https://www.john-fox.ca/Companion/

Gabry, J., & Mahr, T. (2025). bayesplot: Plotting for Bayesian Models (Version R package version 1.15.0) [Computer software]. https://mc-stan.org/bayesplot/

Gabry, J., Simpson, D., Vehtari, A., Betancourt, M., & Gelman, A. (2019). Visualization in Bayesian Workflow. Journal of the Royal Statistical Society Series A: Statistics in Society, 182(2), 389–402. 10.1111/rssa.12378

Gelman, A., Carlin, J. B., Stern, H. S., Dunson, D. B., Vehtari, A., & Rubin, D. B. (2013). Basics of Markov chain simulation. In Bayesian Data Analysis (Third). Chapman & Hall/CRC.

Gohel, D., & Skintzos, P. (2025). flextable: Functions for tabular reporting [Computer software]. 10.32614/CRAN.package.Aextable

Grant, J. E., Odlaug, B. L., & Chamberlain, S. R. (2017). Gambling disorder, DSM-5 criteria and symptom severity. Comprehensive Psychiatry, 75, 1–5. 10.1016/j.comppsych.2017.02.006

Guttman, L. (1950). The basis for scalogram analysis. In S. A. Stouffer, L. Guttman, E. A. Suchman, P. F. Lazarsfeld, S. A. Star, & J. A. Clausen (Eds), Measurement and prediction (pp. 60–90). Princeton University Press.

Hester, J., & Bryan, J. (2024). glue: Interpreted string literals [Computer software]. 10.32614/CRAN.package.glue

Iannone, R., Cheng, J., Schloerke, B., Hughes, E., Lauer, A., Seo, J., Brevoort, K., & Roy, O. (2025). gt: Easily Create Presentation-Ready Display Tables (p. 1.3.0) [Data set]. 10.32614/CRAN.package.gt

Kuznetsova, A., Brockhoff, P. B., & Christensen, R. H. B. (2017). **lmerTest** Package: Tests in Linear Mixed Effects Models. Journal of Statistical Software, 82(13). 10.18637/jss.v082.i13

Lambert, M. J., Morton, J. J., Hatfield, D., Harmon, C., Hamilton, S., Reid, R. C., Shimokawa, K., Christopherson, C., & Burlingame, G. M. (2004). Administration and soring manual for the OQ-45.2 (Outcome Questionnaire). Wilmington DE: American Professional Credential Services.

Lenth, R. V., & Piaskowski, J. (2025). emmeans: Estimated Marginal Means, aka Least-Squares Means (p. 2.0.4) [Data set]. 10.32614/CRAN.package.emmeans

Li, M. F., & Olejnik, S. (1997). The Power of Rasch Person-Fit Statistics in Detecting Unusual Response Patterns. Applied Psychological Measurement, 21(3), 215–231. 10.1177/01466216970213002

Lucas, I., Mora-Maltas, B., Granero, R., Demetrovics, Z., Ciudad-Fernández, V., Nigro, G., Cosenza, M., Rosinska, M., Tapia, J., Fernández-Aranda, F., & Jiménez-Murcia, S. (2024). Network analysis of DSM-5 criteria for gambling disorder: Considering sex differences in a large clinical sample. European Psychiatry, 67(1), e65. 10.1192/j.eurpsy.2024.22

Luo, Y., & Jiao, H. (2018). Using the Stan Program for Bayesian Item Response Theory. Educational and Psychological Measurement, 78(3), 384–408. 10.1177/0013164417693666

Magis, D., Raîche, G., & Béland, S. (2012). A Didactic Presentation of Snijders’s *l_z_ \** Index of Person Fit With Emphasis on Response Model Selection and Ability Estimation. Journal of Educational and Behavioral Statistics, 37(1), 57–81. 10.3102/1076998610396894

Maier, M. J. (2022). Companion Package to the Book ‘R: Einführung durch angewandte Statistik’. https://CRAN.R-project.org/package=REdaS

Masur, P. (2025). ggmirt: Plotting functions to extend ‘mirt’ for IRT analyses [Computer software]. https://github.com/masurp/ggmirt

Meyer, D., Dimitriadou, E., Hornik, K., Weingessel, A., & Leisch, F. (2025). e1071: Misc Functions of the Department of Statistics, Probability Theory Group (Formerly: E1071), TU Wien (p. 1.7-17) [Data set]. 10.32614/CRAN.package.e1071

Meyer, G., Kalke, J., & Buth, S. (2024). Problem gambling in Germany: Results of a mixed-mode population survey in 2021. International Gambling Studies, 24(1), 1–18. 10.1080/14459795.2023.2182337

Mide, M., Arvidson, E., & Gordh, A. S. (2023). Clinical Differences of mild, Moderate, and Severe Gambling Disorder in a Sample of Treatment Seeking Pathological Gamblers in Sweden. Journal of Gambling Studies, 39(3), 1129–1153. 10.1007/s10899-022-10183-x

Mokken, R. J. (1971). A Theory and Procedure of Scale Analysis: With Applications in Political Research. DE GRUYTER MOUTON. 10.1515/9783110813203

Montiel, I., Ortega-Barón, J., Basterra-González, A., González-Cabrera, J., & Machimbarrena, J. M. (2021). Problematic online gambling among adolescents: A systematic review about prevalence and related measurement issues. Journal of Behavioral Addictions, 10(3), 566–586. 10.1556/2006.2021.00055

Moreira, D., Azeredo, A., & Dias, P. (2023). Risk Factors for Gambling Disorder: A Systematic Review. Journal of Gambling Studies, 39(2), 483–511. 10.1007/s10899-023-10195-1

Mousavi, A., Tendeiro, J. N., & Younesi, J. (2016). Person fit assessment using the PerFit package in R. The Quantitative Methods for Psychology, 12(3), 232–242. 10.20982/tqmp.12.3.p232

National Research Council. (1999). Social and economic effects. In Pathological gambling: A critical review (pp. 156–191). National Academies Press.

Nering, M. L., & Meijer, R. R. (1998). A Comparison of the Person Response Function and the lz Person-Fit Statistic. Applied Psychological Measurement, 22(1), 53–69. 10.1177/01466216980221004

Neuwirth, E. (2022). RColorBrewer: ColorBrewer palettes. 10.32614/CRAN.package.RColorBrewer

Orford, J., Wardle, H., Griffiths, M., Sproston, K., & Erens, B. (2010). PGSI and DSM-IV in the 2007 British Gambling Prevalence Survey: Reliability, item response, factor structure and inter-scale agreement. International Gambling Studies, 10(1), 31–44. 10.1080/14459790903567132

Orlando, M., & Thissen, D. (2000). Likelihood-Based Item-Fit Indices for Dichotomous Item Response Theory Models. Applied Psychological Measurement, 24(1), 50–64. 10.1177/01466216000241003

Pedersen, T. L. (2025). patchwork: The Composer of Plots (p. 1.3.2) [Data set]. 10.32614/CRAN.package.patchwork

R Core Team. (2025). R: A Language and Environment for Statistical Computing [Computer software]. R Foundation for Statistical Computing. https://www.R-project.org/

Reeve, B. B., & Fayers, P. M. (2005). Applying item response theory modelling for evaluating questionnaire item and scale properties. In Assessing Quality of Life in Clinical Trials: Methods and Practice 2nd edn (ed Fayers, P. M.; Hays, R. D.) (pp. 55–73). Oxford University Press.

Revelle, W. (2025). psych: Procedures for Psychological, Psychometric, and Personality Research. Northwestern University. https://CRAN.R-project.org/package=psych

Rivera-Riquelme, M., Piqueras, J. A., & Cuijpers, P. (2019). The Revised Mental Health Inventory-5 (MHI-5) as an ultra-brief screening measure of bidimensional mental health in children and adolescents. Psychiatry Research, 274, 247–253. 10.1016/j.psychres.2019.02.045

Rumpf, H.-J., Meyer, C., Hapke, U., & John, U. (2001). Screening for mental health: Validity of the MHI-5 using DSM-IV Axis I psychiatric disorders as gold standard. Psychiatry Research, 105(3), 243–253. 10.1016/S0165-1781(01)00329-8

Sjoberg, D., D., Whiting, K., Curry, M., Lavery, J. A., & Larmarange, J. (2021). Reproducible Summary Tables with the gtsummary Package. The R Journal, 13(1), 570. 10.32614/RJ-2021-053

Sleczka, P., Braun, B., Piontek, D., Bühringer, G., & Kraus, L. (2015). DSM-5 criteria for gambling disorder: Underlying structure and applicability to specific groups of gamblers. Journal of Behavioral Addictions, 4(4), 226–235. 10.1556/2006.4.2015.035

Snijders, T. A. B. (2001). Asymptotic Null Distribution of Person Fit Statistics with Estimated Person Parameter. Psychometrika, 66(3), 331–342. 10.1007/BF02294437

Stan Development Team. (2025). RStan: The R interface to Stan (Version 2.32.7) [Computer software].

Stan Development Team. (n.d.). Item-response theory models. Stan User’s Guide (Version 2.36): Regression Models. https://mc-stan.org/docs/stan-users-guide/regression.html#item-response-models.section

Stinchfield, R. (2002). Reliability, validity, and classification accuracy of the South Oaks Gambling Screen (SOGS). Addictive Behaviors, 27(1), 1–19. 10.1016/S0306-4603(00)00158-1

Stinchfield, R., Govoni, R., & Frisch, G. R. (2005). DSM-IV Diagnostic Criteria for Pathological Gambling: Reliability, Validity, and Classification Accuracy. The American Journal on Addictions, 14(1), 73–82. 10.1080/10550490590899871

Stone, B. M. (2021). The ethic Use of Fit Indices in Structural Equation Modeling: Recommendations for Psychologists. Frontiers in Psychology, 12, 783226. 10.3389/fpsyg.2021.783226

Strand, B. H., Dalgard, O. S., Tambs, K., & Rognerud, M. (2003). Measuring the mental health status of the Norwegian population: A comparison of the instruments SCL-25, SCL-10, SCL-5 and MHI-5 (SF-36). Nordic Journal of Psychiatry, 57(2), 113–118. 10.1080/08039480310000932

Strong, D. R., & Kahler, C. W. (2007). Evaluation of the continuum of gambling problems using the DSM-IV. Addiction, 102(5), 713–721. 10.1111/j.1360-0443.2007.01789.x

Temcheff, C. E., Paskus, T. S., Potenza, Marc. N., & Derevensky, J. L. (2016). Which Diagnostic Criteria are Most Useful in Discriminating Between Social Gamblers and Individuals with Gambling Problems? An Examination of DSM-IV and DSM-5 Criteria. Journal of Gambling Studies, 32(3), 957–968. 10.1007/s10899-015-9591-5

Ten Have, M., Van Bon-Martens, M. J. H., Schouten, F., Van Dorsselaer, S., Shields-Zeeman, L., & Luik, A. I. (2024). Validity of the five-item mental health inventory for screening current mood and anxiety disorders in the general population. International Journal of Methods in Psychiatric Research, 33(3), e2030. 10.1002/mpr.2030

Tendeiro, J. N., Meijer, R. R., & Niessen, A. S. M. (2016). **PerFit**: An *R* Package for Person-Fit Analysis in IRT. Journal of Statistical Software, 74(5). 10.18637/jss.v074.i05

Tierney, N., & Cook, D. (2023). Expanding Tidy Data Principles to Facilitate Missing Data Exploration, Visualization and Assessment of Imputations. Journal of Statistical Software, 105(7). 10.18637/jss.v105.i07

Toce-Gerstein, M., Gerstein, D. R., & Volberg, R. A. (2003). A hierarchy of gambling disorders in the community. Addiction, 98(12), 1661–1672. 10.1111/j.1360-0443.2003.00545.x

Tran, L. T., Wardle, H., Colledge-Frisby, S., Taylor, S., Lynch, M., Rehm, J., Volberg, R., Marionneau, V., Saxena, S., Bunn, C., Farrell, M., & Degenhardt, L. (2024). The prevalence of gambling and problematic gambling: A systematic review and meta-analysis. The Lancet Public Health, 9(8), e594–e613. 10.1016/S2468-2667(24)00126-9

Van Der Flier, H. (1982). Deviant Response Patterns and Comparability of Test Scores. Journal of Cross-Cultural Psychology, 13(3), 267–298. 10.1177/0022002182013003001

Wardle, H., Degenhardt, L., Marionneau, V., Reith, G., Livingstone, C., Sparrow, M., Tran, L. T., Biggar, B., Bunn, C., Farrell, M., Kesaite, V., Poznyak, V., Quan, J., Rehm, J., Rintoul, A., Sharma, M., Shiffman, J., Siste, K., Ukhova, D., … Saxena, S. (2024). The Lancet Public Health Commission on gambling. The Lancet Public Health, 9(11), e950–e994. 10.1016/S2468-2667(24)00167-1

Wickham, H. (2007). Reshaping Data with the **reshape** Package. Journal of Statistical Software, 21(12). 10.18637/jss.v021.i12

Wickham, H., Averick, M., Bryan, J., Chang, W., McGowan, L., François, R., Grolemund, G., Hayes, A., Henry, L., Hester, J., Kuhn, M., Pedersen, T., Miller, E., Bache, S., Müller, K., Ooms, J., Robinson, D., Seidel, D., Spinu, V., … Yutani, H. (2019). Welcome to the Tidyverse. Journal of Open Source Software, 4(43), 1686. 10.21105/joss.01686

Wickham, H., Henry, L., Pedersen, T. L., Luciani, T. J., Decorde, M., & Lise, V. (2025). svglite: An ‘SVG’ Graphics Device (p. 2.2.2) [Data set]. 10.32614/CRAN.package.svglite

Xie, Y. (2014). knitr: A comprehensive tool for reproducible research in R. In V. Stodden, F. Leisch, & R. D. Peng (Eds), Implementing reproducible computational research. Chapman & Hall/CRC.

Xie, Y. (2015). Dynamic documents with R and knitr (Second). Chapman & Hall/CRC. https://yihui.org/knitr/

Xie, Y. (2025). knitr: A general-purpose package for dynamic report generation in R. Chapman & Hall/CRC. https://yihui.org/knitr/

Xie, Y., Allaire, J., & Grolemund, G. (2018). R markdown: The definitive guide. Chapman & Hall/CRC. https://yihui.org/knitr/

Xie, Y., Allaire, J., & Riederer, E. (2020). R markdown cookbook. https://bookdown.org/yihui/rmarkdown-cookbook

Zeileis, A., & Hothorn, T. (2002). Diagnostic checking in regression relationships. R News.

