## Supplementary Materials for "Gambling disorder symptom severity and mental health: an item-response-theory analysis of DSM-5 criteria for gambling disorder"

```
data {
  int<lower=1> J;                        // number of participants
  int<lower=1> K;                        // number of items
  int<lower=1> N;                        // number of observations
  array[N] int<lower=1, upper=J> jj;     // participant for observation n
  array[N] int<lower=1, upper=K> kk;     // item for observation n
  array[N] int<lower=0, upper=1> y;      // correctness for observation n
}

parameters {
  real<lower=-10, upper=10> mu_beta;            // mean item difficulty
  vector[J] alpha;                              // ability for j
  vector[K] beta;                               // difficulty for k
  vector<lower=0>[K] gamma;                     // discrimination of k
  real<lower=0.001, upper=10> sigma_beta;       // scale of difficulties
  real<lower=0.001, upper=10> sigma_gamma;      // scale of log discrimination
}

model {
  alpha ~ std_normal();
  beta ~ normal(0, sigma_beta);
  gamma ~ lognormal(0, sigma_gamma);
  mu_beta ~ uniform(-10, 10);
  sigma_beta ~ uniform(0.001, 10);
  sigma_gamma ~ uniform(0.001, 10);
  y ~ bernoulli_logit(gamma[kk] .* (alpha[jj] - (beta[kk] + mu_beta)));
}
```

**Figure 6.** Stan model specification for the 2PL-IRT model

**Table 4**

*Prevalence of GD in the 2021 and 2023 samples*

| **Prevalence of DSM-5 criteria** | | |
|---|---|---|
| **Criteria** | **2021** ($N_1$ = 1,917) | **2023** ($N_2$ = 979) |
| 1-3 | 1,182 (61.7 %) | 703 (71.8 %) |
| 4-5 | 314 (16.4 %) | 121 (12.4 %) |
| 6-7 | 233 (9.8 %) | 77 (7.9 %) |
| 8-9 | 188 (9.8 %) | 78 (8 %) |

*Note*. Total number (percentage)

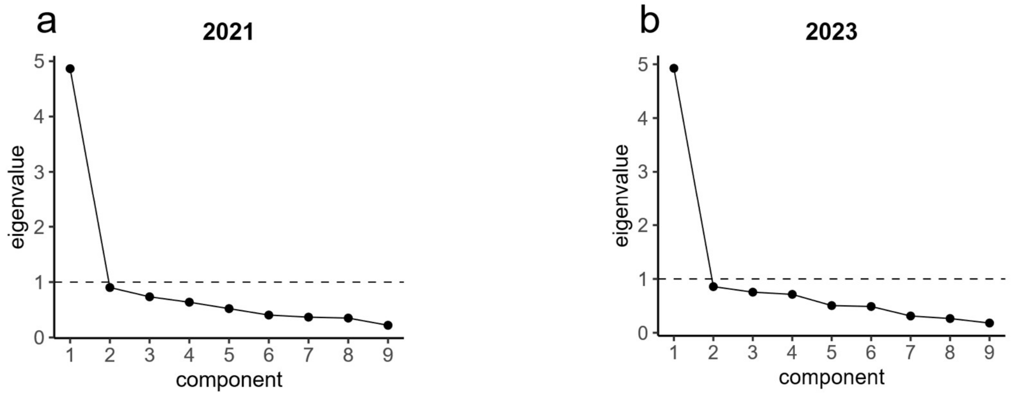


**Figure 7.** Scree plot of eigenvalues for the 2021 sample (**a**) and the 2023 sample (**b**)

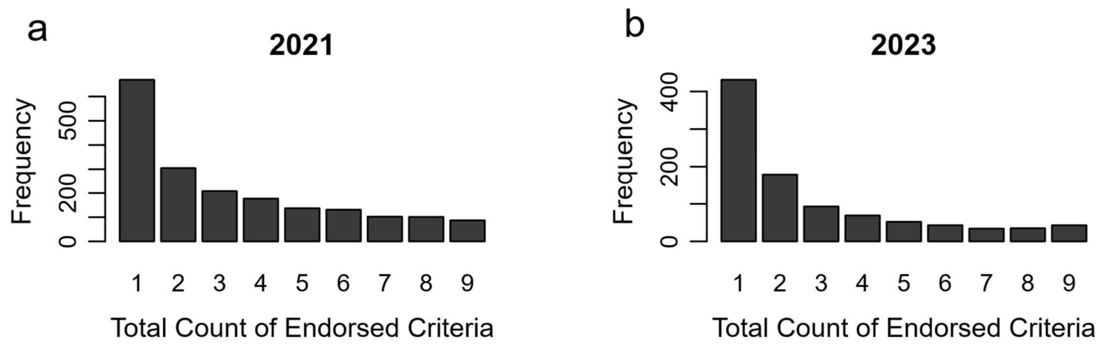


**Figure 8.** Distribution of participants' total counts of endorsed criteria for the 2021 (**a**) and 2023 (**b**) samples

**Table 5**

*Q3 residual correlations for item pairs in the 2021 and 2023 sample*

| **Residual correlations (Yen's Q3)** | | | | | | | | | |
|---|---|---|---|---|---|---|---|---|---|
| **Criteria** | **2021** ($N_1$ = 1,917) | | | | | | | | |
| | DSM5_1 | DSM5_2 | DSM5_3 | DSM5_4 | DSM5_5 | DSM5_6 | DSM5_7 | DSM5_8 | DSM5_9 |
| DSM5_1 | | 0.03 | -0.02 | -0.04 | -0.10 | -0.11 | -0.08 | -0.09 | -0-08 |
| DSM5_2 | 0.03 | | -0.09 | -0.08 | -0.13 | -0.02 | -0.18 | -0.17 | -0.19 |
| DSM5_3 | -0.02 | -0.09 | | 0.01 | -0.09 | -0.10 | -0.14 | -0.19 | -0.13 |
| DSM5_4 | -0-04 | -0-08 | 0.01 | | -0.08 | -0.08 | -0.15 | -0.22 | -0.21 |
| DSM5_5 | -0.10 | -0.13 | -0.09 | -0.08 | | -0.09 | -0.10 | -0.08 | -0.07 |
| DSM5_6 | -0.11 | -0.02 | -0.10 | -0.08 | -0.09 | | -0.07 | -0.09 | -0.11 |
| DSM5_7 | -0.08 | -0.18 | -0.14 | -0.15 | -0.10 | -0.07 | | -0.18 | -0.09 |
| DSM5_8 | -0.09 | -0.17 | -0.19 | -0.22 | -0.08 | -0.09 | -0.18 | | -0.05 |
| DSM5_9 | -0.08 | -0.19 | -0.13 | -0.21 | -0.07 | -0.11 | -0.09 | -0.05 | |
| **Criteria** | **2023** ($N_2$ = 979) | | | | | | | | |
| | DSM5_1 | DSM5_2 | DSM5_3 | DSM5_4 | DSM5_5 | DSM5_6 | DSM5_7 | DSM5_8 | DSM5_9 |
| DSM5_1 | | 0.00 | -0.05 | -0.02 | -0.07 | -0.08 | -0.12 | -0.10 | -0.05 |
| DSM5_2 | 0.00 | | -0.08 | -0.14 | -0.12 | -0.03 | -0.19 | -0.15 | -0.10 |
| DSM5_3 | -0.05 | -0.08 | | 0.00 | -0.14 | -0.11 | -0.10 | -0.13 | -0.14 |
| DSM5_4 | -0.02 | -0.14 | 0.00 | | -0.07 | -0.03 | -0.16 | -0.21 | -0.25 |
| DSM5_5 | -0.07 | -0.12 | -0.14 | -0.07 | | -0.11 | -0.09 | -0.08 | -0.06 |
| DSM5_6 | -0.08 | -0.03 | -0.11 | -0.03 | -0.11 | | -0.11 | -0.10 | -0.07 |
| DSM5_7 | -0.12 | -0.19 | -0.10 | -0.16 | -0.09 | -0.11 | | -0.09 | -0.09 |
| DSM5_8 | -0.10 | -0.15 | -0.13 | -0.21 | -0.08 | -0.10 | -0.09 | | -0.10 |
| DSM5_9 | -0.05 | -0.10 | -0.14 | -0.25 | -0.06 | -0.07 | -0.09 | -0.10 | |

*Note.* Residual correlations were compared against a cut-off score of $|Q3| < 0.2$. Residual correlations between criteria 4 and 8 as well as 4 and 9 slightly exceed the cut-off in both samples.

**Table 6**

*Loadings and communalities of the one-factor model for both samples*

| **One-Factor Model: Loadings and Communalities** | | | | |
|---|---|---|---|---|
| **Criteria** | **2021** ($N_1$ = 1,917) | | **2023** ($N_2$ = 979) | |
| | Loading (λ) | Communality ($h^2$) | Loading (λ) | Communality ($h^2$) |
| DSM5_1 | 0.46 | 0.21 | 0.47 | 0.22 |
| DSM5_2 | 0.69 | 0.47 | 0.68 | 0.47 |
| DSM5_3 | 0.64 | 0.40 | 0.61 | 0.38 |
| DSM5_4 | 0.76 | 0.58 | 0.82 | 0.67 |
| DSM5_5 | 0.62 | 0.38 | 0.64 | 0.41 |
| DSM5_6 | 0.56 | 0.31 | 0.55 | 0.31 |
| DSM5_7 | 0.78 | 0.60 | 0.78 | 0.61 |
| DSM5_8 | 0.87 | 0.76 | 0.85 | 0.72 |
| DSM5_9 | 0.82 | 0.68 | 0.85 | 0.72 |

*Note*. Loadings (λ) from ML one-factor EFA; $h^2$ = loading$^2$

**Table 7**

*Infit and outfit statistics for 2PL-IRT models of the 2021 and 2023 sample using the mirt package*

| **Criteria** | **2021** ($N_1$ = 1,917) | | **2023** ($N_2$ = 979) | |
|---|---|---|---|---|
| | Infit | Outfit | Infit | Outfit |
| DSM5_1 | 0.95 | 0.92 | 0.94 | 0.93 |
| DSM5_2 | 0.85 | 0.74 | 0.87 | 0.76 |
| DSM5_3 | 0.87 | 0.82 | 0.87 | 0.83 |
| DSM5_4 | 0.92 | 0.68 | 0.96 | 0.60 |
| DSM5_5 | 0.90 | 0.86 | 0.89 | 0.85 |
| DSM5_6 | 0.89 | 0.85 | 0.90 | 0.87 |
| DSM5_7 | 0.86 | 0.70 | 0.89 | 0.74 |
| DSM5_8 | 0.84 | 0.58 | 0.93 | 0.66 |
| DSM5_9 | 0.87 | 0.73 | 0.92 | 0.69 |

**Table 8**

*Medians and 95% credible intervals of 2PL-IRT item parameters for the 2021 and 2023 sample*

| **Criteria** | **2021** ($N_1$ = 1,917) | | **2023** ($N_2$ = 979) | |
|---|---|---|---|---|
| | Discrimination (gamma) | Difficulty (beta) | Discrimination (gamma) | Difficulty (beta) |
| DSM5_1 | 0.71 [0.58, 0.85] | -0.61 [-1.13, -0.12] | 0.68 [0.50, 0.88] | 0.03 [-0.46, 0.47] |
| DSM5_2 | 1.61 [1.41, 1.82] | 0.28 [-0.22, 0.76] | 1.46 [1.19, 1.77] | 0.60 [0.14, 1.03] |
| DSM5_3 | 1.32 [1.14, 1.50] | 0.24 [-0.25, 0.73] | 1.16 [0.93, 1.42] | 0.37 [-0.10, 0.79] |
| DSM5_4 | 1.89 [1.65, 2.16] | 1.10 [0.61, 1.59] | 2.11 [1.72, 2.57] | 1.25 [0.80, 1.69] |
| DSM5_5 | 1.16 [1.01, 1.33] | 0.38 [-0.12, 0.86] | 1.14 [0.92, 1.39] | 0.60 [0.14, 1.03] |
| DSM5_6 | 1.06 [0.90, 1.23] | -0.16 [-0.65, 0.33] | 0.93 [0.72, 1.15] | 0.15 [-0.33, 0.57] |
| DSM5_7 | 1.89 [1.66, 2.15] | 0.59 [0.10, 1.08] | 1.76 [1.45, 2.13] | 0.89 [0.44, 1.33] |
| DSM5_8 | 2.52 [2.19, 2.91] | 0.94 [0.44, 1.42] | 2.13 [1.75, 2.61] | 1.23 [0.77, 1.66] |
| DSM5_9 | 2.11 [1.84, 2.42] | 1.17 [0.68, 1.66] | 2.13 [1.74, 2.60] | 1.37 [0.92, 1.83] |

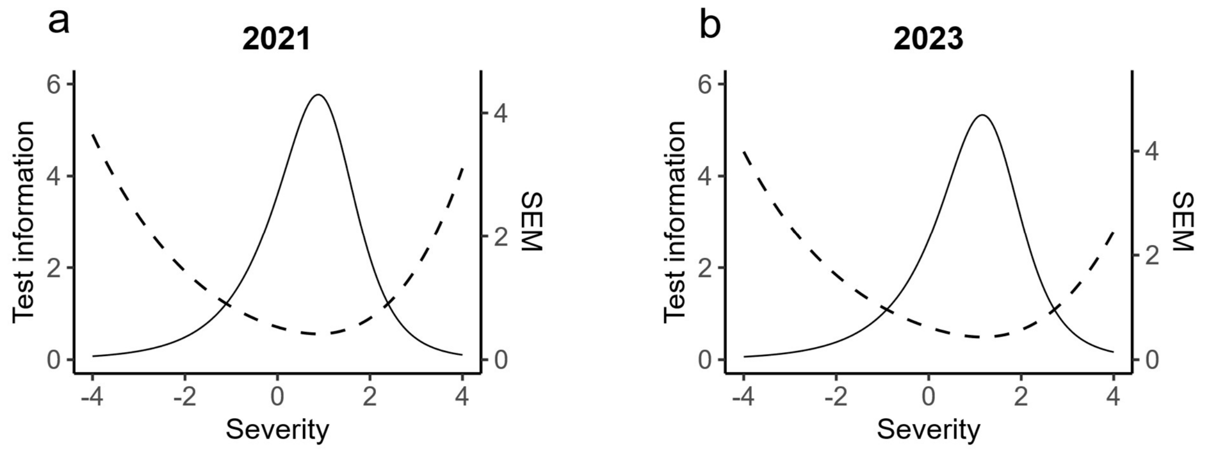


**Figure 9.** Total information functions (TIF) for the 2021 (**a**) and 2023 (**b**) sample

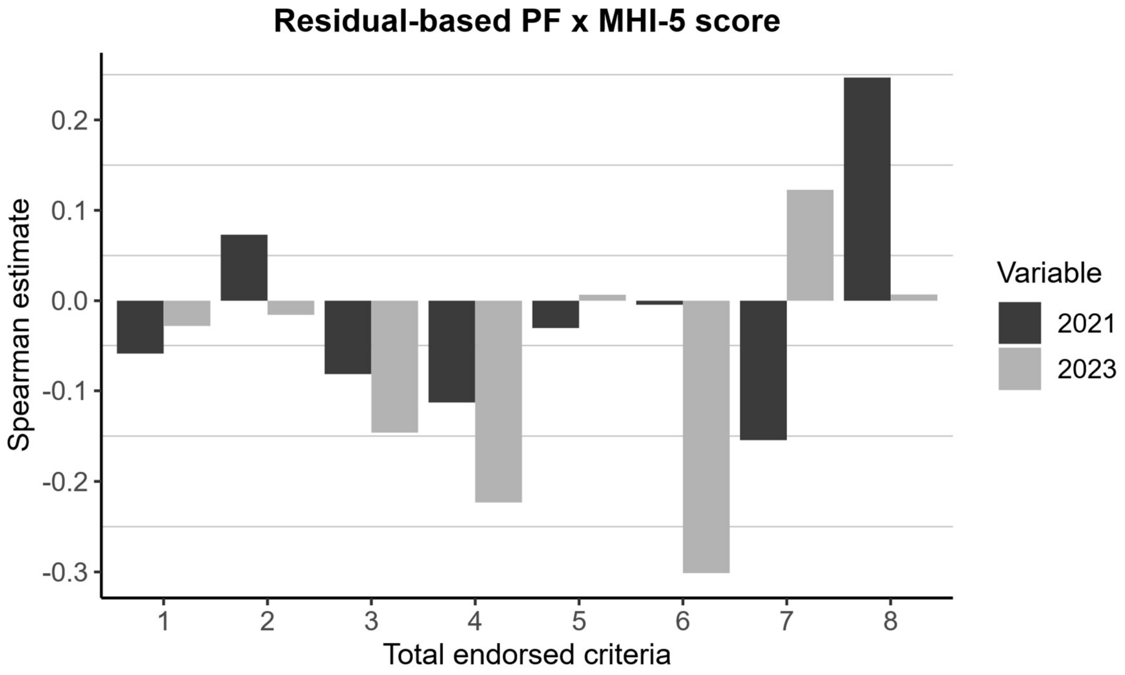


**Figure 10.** Correlations between residual-based PF and MHI-5 scores for groups of total endorsed criteria of each sample

**Table 9**

*Random intercept model for residual-based PF index*

| **Statistic** | **Values** |
|---|---|
| Slope (β) | -0.57 |
| SE | 0.35 |
| t(df) | -1.63 (2873.2) |
| p | 0.10 |
| SD(group) | 5.48 |
| SD(sample) | 0.48 |
| SD(residual) | 17.93 |
| ICC(group) | 0.09 |
| ICC(sample) | 0.00 |

B = fixed PF slope (within total-score groups); p via Scatterthwaite df.
ICC(group) = Var(group)/[Var(group)+Var(sample);
ICC(sample) = Var(sample)/[Var(group)+Var(sample)+Var(residual)].
